# Evaluating Wastewater Surveillance for Estimating Pharmaceutical Use

**DOI:** 10.1101/2025.05.16.25327761

**Authors:** Gilbert Osena, Jerker Fick, Carl-Fredrik Flach, Erik Kristiansson, D. G. Joakim Larsson

## Abstract

Regional medicine use data is key for health management and in understanding many aspects of health, still such data is missing in many parts of the world. Here, we aimed to evaluate how well analyses of active pharmaceutical ingredients (APIs) in untreated municipal wastewater could be used to predict regional use. We studied 53 APIs measured at three wastewater treatment plants (WWTPs) in Stockholm, Sweden between 2004-2021 together with corresponding, comprehensive sales data from hospitals, pharmacies and other outlets as proxy for use. Conversion factors (CFs) representing recovered mass per gram of sold API were generated for each API using data from two WWTPs. Median absolute deviation normalized by the median (MADM) was used to evaluate variability of CFs over the years. While CFs ranged more than two orders of magnitude between different APIs, mass loads recovered in wastewater were equal or less than the estimated daily use for all except one API. The variability of CF estimates was below 100% for 43 APIs. When evaluating the predictive performance of the derived CFs on data from the third WWTP, the typical error was <2-fold for 36 APIs (68%). Neither removal efficiency in WWTPs nor lipophilicity were good predictors of CFs. Our findings suggest that use of most APIs can be estimated from traces measured in wastewater with a typical error of <2-fold. This provides support for the development of wastewater surveillance for estimating medicine use to fill existing data gaps, but also suggest limitations in detecting small changes.

## 1. Introduction

Consumption data for medicines is not known or highly uncertain in many parts of the world, mainly due to inefficient systems to track sales and due to unregulated access. Reliable consumption data for pharmaceuticals in a population provides insights into its health profile, and/or if there seems to be an under- or over-use of certain medicines. Thus, data on pharmaceutical use facilitates evaluation of needs as well as of impacts of policies affecting use, including prescription recommendations, distribution and pricing of targeted medicines (WHO, 1993).

One should acknowledge that even the most accurate sales data will only be a proxy for use. Unregulated/unaccounted sales will lead to underestimations of use, whereas large proportions of sold but discarded medicines will lead to overestimations (Kasprzyk-Hordern et al., 2021). Conventional approaches of estimating pharmaceutical use data involve collating prescription data, or sales data from pharmacies, pharmaceutical suppliers and manufacturers (Escolà Casas et al., 2021). However, accessing the data by either of these approaches can be difficult due to privacy issues with prescription data, and expensive due to the manual work involved or the potential commercial value of sales data. In some settings this limitation is overcome by automated systems that record prescription and sales data of pharmaceuticals in centralized databases. Without such systems, pharmaceutical use data are usually scattered in multiple health facilities and pharmacy records needing much effort to collect, which could easily underestimate use due to missed sources. Therefore, alternative approaches that could measure pharmaceutical use are desirable to complement the existing ones.

Environmental monitoring has increasingly been recognized as a way to generate data that reflects societal activities, not least in relation to human health. In particular, wastewater-based epidemiology (WBE) has been applied in many parts of the world to track disease prevalences and outbreaks, monitor human exposure to chemicals in the environment, and to monitor illicit drug use and alcohol consumption (Been et al., 2018; Bijlsma et al., 2016; López-García et al., 2020; Saguti et al., 2021). To estimate pharmaceutical drug consumption, wastewater surveillance relies on the assumption that a relatively stable proportion of the active pharmaceutical ingredients (APIs) eventually end up in wastewater after use and excretion. A few studies have reported back-calculated pharmaceutical drug consumption estimates from the concentrations of APIs measured in untreated influents from the inlets of wastewater treatment plants (WWTPs) (Escolà Casas et al., 2021; Gao et al., 2021; van Nuijs et al., 2015). These studies, however, do not always report consistent conversion factors (CFs) for the same APIs, and the number of investigated APIs is limited, as is replication of estimates over time and space. A CF of an API is obtained by dividing mass loads recovered in untreated wastewater with the corresponding estimated use and could be used to back-calculate usage of the parent drug in the catchment population. The generation of CFs is therefore dependent on good sales/usage data in the studied catchment. Once reliable and robust CFs are generated, they would enable estimates of medicine use in settings lacking good quality sales data. Some of the factors that could contribute to the variability of CF estimates include the quality and spatio-temporal resolution of pharmaceutical use data, differences in the studied populations with regards to e.g. drug metabolism, differences in the sewers systems that may affect the fate of APIs, methods used for wastewater sampling and chemical analysis, as well as different approaches for back calculation. The underlying assumption in estimating conversion factors is that there is a negligible proportion of unaccounted sales (such as for illegal use), and that all purchased drugs are used by the patients and later excreted into the sewer systems. However, previous studies have reported that 3 – 50% of all prescribed drugs are not used by the patients (Boxall et al., 2014; OECD, 2022), which could lead to underestimations of the CFs. Further, 0 - 54% of respondents in different studies have admitted to flushing unused medicines in the toilets (Persson et al., 2009; Rogowska and Zimmermann, 2022; Seehusen and Edwards, 2006; Tong et al., 2011) which could lead to over-estimation of the conversion factors. High quality reference data on pharmaceutical use together with API wastewater surveillance data covering multiple time points and many APIs with different characteristics could help in estimating uncertainties around the CFs, and to evaluate the overall potential to analyze medicines in wastewater to assess regional usage. Sweden has an excellent and long-standing record of medicine use (Furu et al., 2010), and low levels of non-regulated use for most pharmaceuticals (Läkemedelsverket, 2023). Besides, in surveys conducted in Sweden between 2001 and 2007, no respondents declared that they disposed of any of their unused medicines into the toilets. On a more hypothetical question on what they would do if they had leftover medicines, only 1-2% responded that they would flush unused medicines. These percentages are lower than in several other surveys in other countries (Castensson and Ekedahl, 2010). This likely stems from the introduction of a nationwide system in Sweden already in 1971 to return unused drugs to pharmacies (Persson et al., 2009), and that Swedes tend to have a relatively high trust in authorities and to follow recommendations to a greater extent than people from many other countries, as demonstrated for example during the Covid-19 pandemic (Kavaliunas et al., 2020). Because of the accessibility to high-quality sales data, demonstrated minimal practice of flushing unused drugs and, in an international context, limited unaccounted, illegal sales, the Swedish setting provides very good possibilities for evaluating a wastewater-based surveillance approach.

Between 2004 and 2021 (18 years), the Stockholm Region, formerly “Stockholms Läns Landsting”, monitored API concentrations in influent and effluent samples collected from the three largest WWTPs (Henriksdal, Käppala and Bromma) in Stockholm. The intended use of the collected chemical analyses data was to assess environmental risks, efficiency of the WWTPs in removing APIs, and determine whether interventions on drug use could be reflected by amounts measured in wastewaters and the environment. However, for the present study, the dataset provided an excellent opportunity to estimate CFs for the measured APIs and their variability.

The aim of the present study was to generate CFs for many APIs and to assess the variability of CF estimates over time, and investigate the impact of different inherent characteristics of the APIs, such as lipophilicity and their persistence to removal within WWTPs on CFs. To achieve this, we took advantage of pharmaceutical sales data and chemical analyses data for 53 APIs in three different WWTPs over 18 years in Stockholm.

## 2. Methods

### 2.1 Study sites

We base the study on analyses of pharmaceutical residues between 2004 and 2021 in wastewater from three WWTPs in Stockholm, Sweden, namely, Henriksdal (59°32’29“N 18°08’30”E), Käppala (59°35’04“N 18°10’26”E), and Bromma (59°31’03“N 17°53’20”E). In 2022, Stockholm had a population of ∼2.4 million, of which approximately 878,000 people (36.0%) were connected to the Henriksdal WWTP, 500,000 (20.5%) to the Käppala WWTP, and 379,800 people (15.6%) to the Bromma WWTP. These three treatment plants serve about 70% of Stockholm’s population. While there has been some (limited) population growth in the catchment area over the 18 years, we have assumed that the relative proportion contributing to each of the three sites has remained constant. From each WWTP, we obtained average daily flow data (m^3^/s) on the sampling days.

### 2.2 Levels of APIs in wastewater

Concentrations (ng/L) of active pharmaceutical ingredients (APIs) measured in samples collected at the inlets and outlets of three wastewater treatment plants (WWTPs) in the Stockholm region were obtained for the period between 2004 and 2021. Each year, sampling was done in August or September, and each sample was a flow-proportional composite collected at each site over 24 hours. Concentrations below the reported limit of quantification (LoQ) were excluded from the comparison with sales data.

Detailed information on how the chemical analyses were performed between 2004 and 2011 has not been possible to retrieve. Briefly though, the analyses were pretreated using solid phase extraction and analyzed with Liquid chromatography coupled with tandem mass spectrometry.

Between 2012 and 2021, the analyses were performed by Umeå University, Sweden. Here the wastewater samples were pretreated using solid phase extraction (Oasis HLB, 6 CC) after adding 50ng of each internal surrogate standard. Samples were then analyzed using a system with a triple-stage quadrupole mass spectrometer (Quantum Ultra EMR (Thermo Fisher Scientific, San Jose, CA) coupled with a liquid chromatographic pump (Accela, Thermo Fisher Scientific) and an autosampler (PAL HTC, CTC Analytics AG, Zwingen, Switzerland). Heated electrospray (HESI), krypton 10.6 eV, in positive ion mode was used for ionization of the pharmaceuticals. Specific details related to the determination of the pharmaceuticals including HESI ionizations, polarities, precursor/product ions, collision energies, tube lens values, etc. have been described in detail elsewhere (Grabic et al., 2012; Lindberg et al., 2014). To ensure proper quality assurance and quality control, two MS/MS transitions were used for positive identifications of analytes with the criterion that the ratio between the transitions was not allowed to deviate more than +/−30% from the ratio in the corresponding calibration standard. Retention times for all analytes also had to be within +/−2.5% of the retention time in the corresponding calibration standard. The limit of quantification (LOQ) was determined from standard curves based on repeated measurements of low level spiked plasma and tissue samples, and the lowest point in the standard curve that had a signal/noise ratio of 10 was considered to be equal to the LOQ. Samples were quantified using a seven-point matrix adjusted calibration curve over the range of 0.05–100 ng ml^-1^. Carry-over effects were evaluated by injecting standards at 100 ng l^−1^ followed by two mobile phase blanks. Several instrumental and field blanks were included in each analytical run. This methodology did not change over the time period of 2012-2021, however additional internal standards were added over the years (Table 1).

**Table 1:** Estimates of conversion factors (proportion recovered in untreated sewage in relation to sales; CFs) and removal efficiencies of 53 active pharmaceutical ingredients (APIs) measured at three wastewater treatment plants in Stockholm, Sweden 2004-2021.

| Name | Year isotope labelled standard was introduced in lab* | Log P | Conversion factor (95% CI) | Removal efficiency (%) (95% CI) |
| --- | --- | --- | --- | --- |
| Alfuzosin |  | 1.9 | 0.208 (0.119 - 0.297) | 1.5 (-29.4 - 32.5) |
| Amlodipine |  | 2.1 | 0.072 (0.052 - 0.093) | 70.6 (52.2 - 89) |
| Atenolol |  | 0 | 0.609 (0.458 - 0.76) | 50.8 (42.5 - 59) |
| Atorvastatin |  | 5.39 | 0.107 (0.089 - 0.125) | 94.8 (90.9 - 98.6) |
| Bisoprolol |  | 1.8 | 0.299 (0.129 - 0.47) | 34.1 (15.8 - 52.4) |
| Bupropion |  | 3.9 | 0.015 (0.008 - 0.023) | 9.6 (-22.3 - 41.4) |
| Carbamazepine | 2012 | 2.2 | 0.06 (0.044 - 0.076) | -27.1 (-42 - -12.2) |
| Ciprofloxacin | 2012 | 0 | 0.121 (0.075 - 0.167) | 86.4 (81.5 - 91.2) |
| Citalopram |  | 3.7 | 0.127 (0.098 - 0.155) | 6.2 (-18.7 - 31) |
| Clarithromycin |  | 3.2 | 0.2 (0.11 - 0.289) | 28.5 (0.7 - 56.3) |
| Clindamycin | 2014 | 2 | 0.016 (0.004 - 0.028) | -125.3 (-229.1 - -21.5) |
| Codeine | 2012 | 1.3 | 0.349 (0.277 - 0.421) | 76 (70.3 - 81.7) |
| Cyclophosphamide |  | 0.76 | 0.593 (0.353 - 0.832) | 55.4 (40.7 - 70.1) |
| Diclofenac | 2012 | 4 | 0.098 (0.057 - 0.14) | 21.9 (12.9 - 31) |
| Diltiazem |  | 2.8 | 0.024 (0.007 - 0.04) | 41.7 (22.9 - 60.4) |
| Dipyridamole |  | 2.7 | 0.699 (0.468 - 0.93) | - |
| Enalapril |  | -0.9 | 0.136 (0.1 - 0.171) | - |
| Eprosartan |  | 3.75 | 1.152 (0.664 - 1.641) | 49.2 (34.7 - 63.8) |
| Erythromycin | 2012 | 2.6 | 0.199 (0.105 - 0.294) | 31.2 (17.6 - 44.9) |
| Felodipine |  | 4.5 | 0.062 (0.05 - 0.075) | 30.9 (15.9 - 45.9) |
| Fexofenadine |  | 2.8 | 0.748 (0.211 - 1.285) | 52.5 (27.8 - 77.2) |
| Flecainide | 2012 | 4 | 0.281 (0.213 - 0.348) | -9 (-27.2 - 9.3) |
| Fluconazole | 2012 | 0.3 | 0.418 (0.256 - 0.58) | 3.5 (-14 - 21) |
| Fluoxetine | 2012 | 4.7 | 0.06 (0.041 - 0.079) | 22.2 (-6.2 - 50.7) |
| Furosemide |  | 2.3 | 0.43 (0.319 - 0.54) | 11 (-5 - 27.1) |
| Hydrochlorothiazide |  | -0.1 | 0.53 (0.38 - 0.681) | -14.3 (-38.2 - 9.5) |
| Ibuprofen | 2012 | 3.8 | 0.101 (0.057 - 0.146) | 99.6 (98.9 - 100.2) |
| Ketoprofen | 2012 | 3 | 0.281 (0.15 - 0.411) | 70 (61.2 - 78.8) |
| Losartan |  | 4 | 0.429 (0.305 - 0.552) | 65.5 (58.1 - 72.8) |
| Memantine |  | 3.3 | 0.208 (0.076 - 0.341) | 24.3 (6.8 - 41.8) |
| Metoprolol | 2019 | 1.7 | 0.094 (0.072 - 0.115) | -1.6 (-16 - 12.8) |
| Metronidazole |  | 0 | 0.036 (0.021 - 0.051) | 52.8 (43.4 - 62.2) |
| Mirtazapine |  | 3 | 0.076 (0.048 - 0.103) | 22.7 (6 - 39.4) |
| Naproxen | 2012 | 3.1 | 0.194 (0.09 - 0.298) | 93.3 (87.6 - 99) |
| Norfloxacin |  | -0.3 | 0.211 (0.105 - 0.316) | - |
| Ofloxacin | 2012 | -0.2 | 3.593 (1.05 - 6.136) | 70 (58.6 - 81.3) |
| Orphenadrine (Citate) |  | 3.7 | 0.442 (0.111 - 0.772) | 38.1 (19.3 - 56.9) |
| Oxazepam | 2012 | 2.3 | 0.609 (0.504 - 0.714) | -5.9 (-16.2 - 4.4) |
| Paracetamol | 2012 | 0.3 | 0.102 (0.048 - 0.156) | 99.8 (99.6 - 100) |
| Propranolol |  | 2.6 | 0.135 (0.099 - 0.17) | 2.6 (-20.9 - 26) |
| Ramipril |  | -0.1 | 0.074 (0.018 - 0.129) | - |
| Ranitidine |  | 0.3 | 0.052 (0.022 - 0.082) | 35.7 (15.8 - 55.6) |
| Rosuvastatin |  | 2.5 | 0.728 (0.554 - 0.903) | 82.1 (76 - 88.1) |
| Sertraline |  | 5.3 | 0.013 (0.007 - 0.019) | 26 (-10 - 62) |
| Sotalol |  | 0.4 | 0.488 (0.331 - 0.646) | 45.9 (25.8 - 66) |
| Telmisartan |  | 6.13 | 1.121 (0.552 - 1.691) | - |
| Terbutaline |  | 0.7 | 0.369 (0.24 - 0.498) | 31.2 (14.3 - 48.2) |
| Tetracycline | 2014 | -1.3 | 0.705 (0.382 - 1.028) | - |
| Tramadol | 2012 | 3 | 0.142 (0.07 - 0.214) | -11.8 (-26.9 - 3.4) |
| Trimethoprim | 2012 | 0.7 | 0.183 (0.135 - 0.23) | 53.9 (41.9 - 65.9) |
| Warfarin |  | 2.2 | 0.052 (0.038 - 0.066) | 0.3 (-33.6 - 34.2) |
| Zolpidem |  | 3.8 | 0.013 (0.006 - 0.021) | -0.7 (-27 - 25.6) |
| Zopiclone |  | 1.5 | 0.127 (0.081 - 0.173) | 18.6 (-8.5 - 45.7) |

‘\*’ : Laboratory at Umeå university that measured active pharmaceutical ingredients in wastewater samples between 2012-2021. ‘-’; Not available due to insufficient data.

### 2.3 Sales data of APIs

August and September sales data of pharmaceuticals in the Stockholm region were retrieved from a central database for the period spanning 2004 to 2021, kindly compiled and provided by Region Stockholm for our specific objectives. The four types of sales captured in the database included over-the-counter sales, open care requisitions, prescriptions, and hospital requisitions. Each sales record included information on the product number and name, anatomical therapeutic chemical (ATC) code and name, type of sale, units or packages sold, and the total defined daily doses (DDDs) sold within the month.

To convert the total DDDs sold for each API to mass units e.g., grams, we retrieved the information of the respective DDD values, units, and strength(s) of the APIs, from each product’s description in the pharmaceuticals’ sales database. The retrieved DDD or unit dose (UD) information of the pharmaceutical products was then used to calculate the sold API masses by multiplying by the tally of DDDs sold. For a drug with a single API, the number of DDDs sold was multiplied by its DDD to obtain the mass sold and converted to grams based on the initial DDD unit (micrograms, milligrams or milliliters). For a drug with more than one API, the quantities were calculated by multiplying the number of DDDs sold by its unit dose (UD) and the strength of each API. These were then converted to grams depending on the units of strength. Where information on DDDs was unavailable (such as most gels), we searched through Swedish drug information databases, including the SIL (www.silonline.silinfo.se) and the FASS databases (www.fass.se) for the package sizes and mass units. For such products with no DDD available, the mass sold was calculated by multiplying the number of units sold by the respective masses.

### 2.4 Calculation of CFs

Given that the sales data were for the entire Stockholm region, we estimated API usage for each WWTP catchment population by multiplying the total grams sold by the respective population proportions as indicated above. Daily usage estimates were obtained through division by 61 (the total number of days in August and September). The quantities of APIs recovered in wastewater from the 24-hour composite samples were calculated as a product of the measured concentration (ng/L) and mass flow rate (m^3^/s) of each of the influents on each sampling day (Supplementary file 1). For each API, we estimated the fraction recovered in wastewater for each gram estimated to be used in the respective catchment. Henceforth, we refer to this metric as the conversion factor (CF). We generated the CFs using data from two sites (Henriksdal and Käppala WWTPs) and used data from Bromma to evaluate the errors associated with applying the CFs to API mass loads recovered in the wastewater to back calculate usage. We decided to hold out Bromma data because it was the median based on the number of all measurements across all APIs (Henriksdal – 664, Bromma – 663, Käppala - 653). The recovered API masses from Bromma were divided by the respective CFs to estimate usage. Mean absolute errors between the log-transformed reported and predicted usage values were calculated for each API and translated to a non-log scale to express the errors in fold -change units. In calculating the CF, we assume that all sold APIs are used and eventually contributing to the excreted amount in the sewers. This assumption should be taken into account when applying CF in other context to predict either sales or use. Due to the presence of outliers and skewedness in the CFs distributions for most APIs over the study period, we used the median measure to represent the CF for each API. APIs with both sales and wastewater data covering at least six years were used in further analysis.

### 2.5 Lipophilicity and removal efficiency

One hypothesis was that characteristics of the APIs could influence the corresponding CFs, including their variability. Data on lipophilicity (logP), which can play an important role in the fate and distribution of APIs between water and solid phases, was primarily sourced from (Fick et al., 2010) and where not available the ALOGPs logP estimate from www.drugbank.com (Knox et al., 2024) was then used. The persistence of the API for removal within WWTPs was also hypothesized to influence CFs and CF-variability. As a measure of persistence, we used the median of the measured removal efficiency of each API in the studied WWTPs. Removal efficiency was obtained by dividing the difference between influent and corresponding effluents by the influent concentrations ((Influent – Effluent)/Influent) and expressed as a percentage. Confidence intervals (95%) were constructed for each API’s removal efficiency and CF based on the standard error of the median. The variabilities around removal efficiencies and CFs were evaluated using the median absolute deviation (MAD) of site-level data per API across the study period, divided by the respective medians and expressed as a percentage (MADM).

### 2.6 Statistical analysis

We used simple linear regression to evaluate relationships between (i) CFs and lipophilicity, (ii) removal efficiency and lipophilicity, and (iii) CFs and removal efficiency. The CFs were log-transformed before the regression analyses to improve the normality of residuals. Wilcoxon rank sum test was used to compare CFs between two periods (2004-2011, and 2012-2021) following the change of contracted analytical laboratories in 2012. Comparisons were performed for drugs with at least six data points in each period, and the false discovery rate (FDR) method was used to correct for multiple hypothesis testing. To compare variability of the CFs between periods based on testing laboratories (2004-2011, and 2012-2021) and whether isotope-labelled standards were used in respective chemical analyses, we used the Wilcoxon signed rank test. Statistical significance was set at p<0.05, and all statistical analyses were conducted in the software R.

## 3. Results

In total, 109 APIs were measured in wastewater influent samples at least once between 2004 and 2021. The sales data, used here as a close proxy for usage, varied over time for the different APIs. We included 53 APIs (Supplementary Figures 1 and 2) in the analysis after removing 55 that did not fulfil the criterion of having both sales and wastewater data for at least six years. Ketoconazole was also excluded, because in all instances when it was measured in the WWTPs, the quantity recovered was 2 - 33 times (median: 8.6) higher than that sold in the healthcare facilities and pharmacies, implying a possible major external ketoconazole source not captured in the sales database. As an example, the estimated daily usage of trimethoprim and recovery in wastewater over the study period is shown in Figure 1, and the other studied APIs in Supplementary Figure 3.

**Figure 1.**
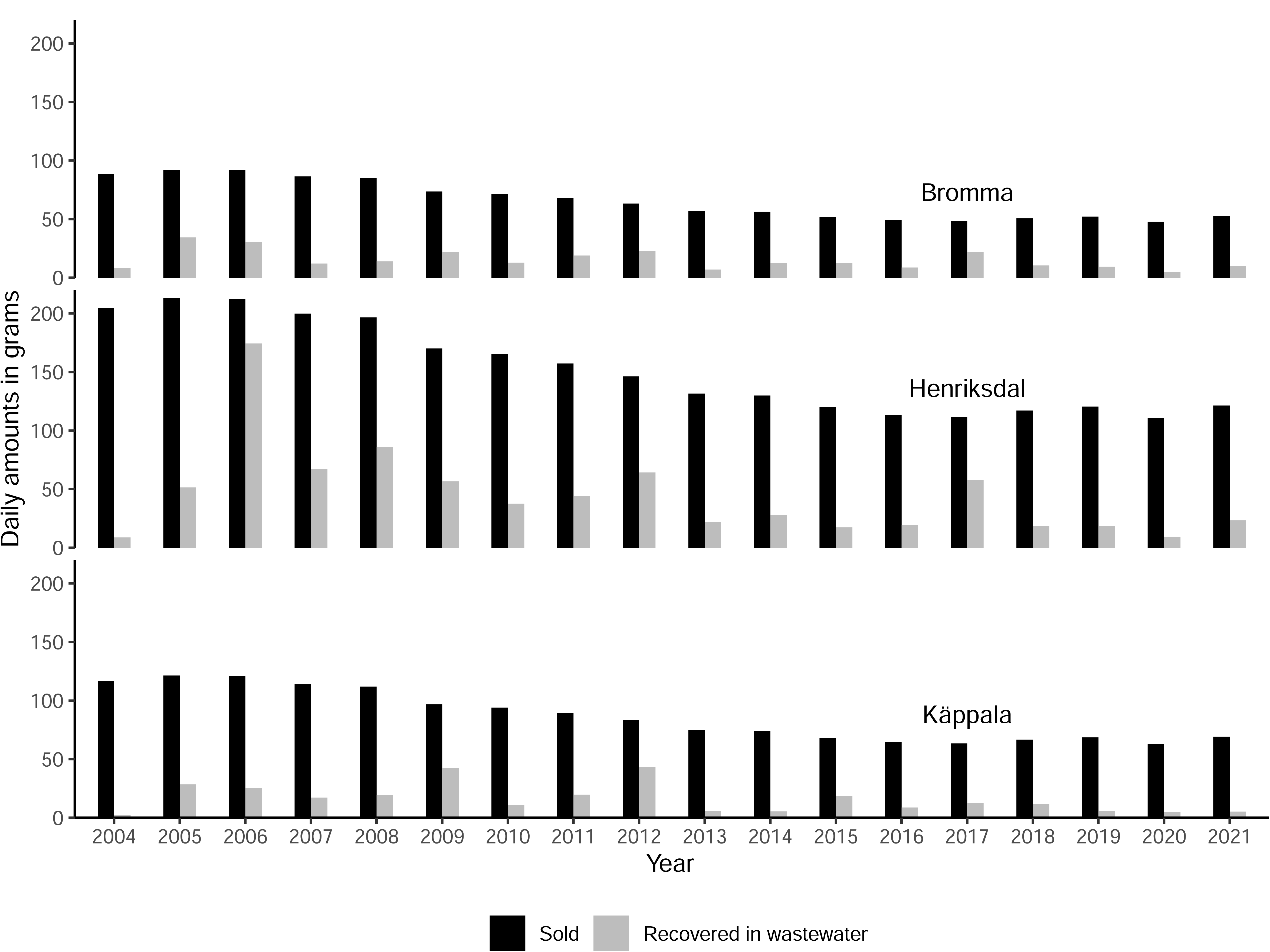
Estimated quantities of trimethoprim sales (black bars) in catchment areas of three wastewater treatment plants (WWTPs) in Stockholm, Sweden and the corresponding recovered masses (grey bars) in WWTPs influents over 24h in August/September 2004-2021. Dark and light-grey bars represent quantities sold and the quantities recovered in the respective WWTP.

Our estimates of fractions of APIs recovered in wastewater per gram used (CF) ranged from 0.013 for sertraline (95% CI 0.006 - 0.021) and zolpidem (95% CI 0.007 - 0.019) to 3.593 for ofloxacin (3.593, 1.05 – 6.136) (Table 1; Figure 2). For 50 (94.3%) APIs, the quantities recovered in wastewater were less than the estimated daily use. For ofloxacin (3.59g), eprosartan (1.15g), and telmisartan (1.12g), the median quantities recovered in wastewater were more than 1 gram per estimated used gram of API, but for eprosartan and telmisartan the 95% confidence interval also included values below 1 gram. Therefore, based on the 95% CI limits, quantities recovered in wastewater were less than the estimated daily use for 52 APIs, except for ofloxacin.

**Figure 2:**
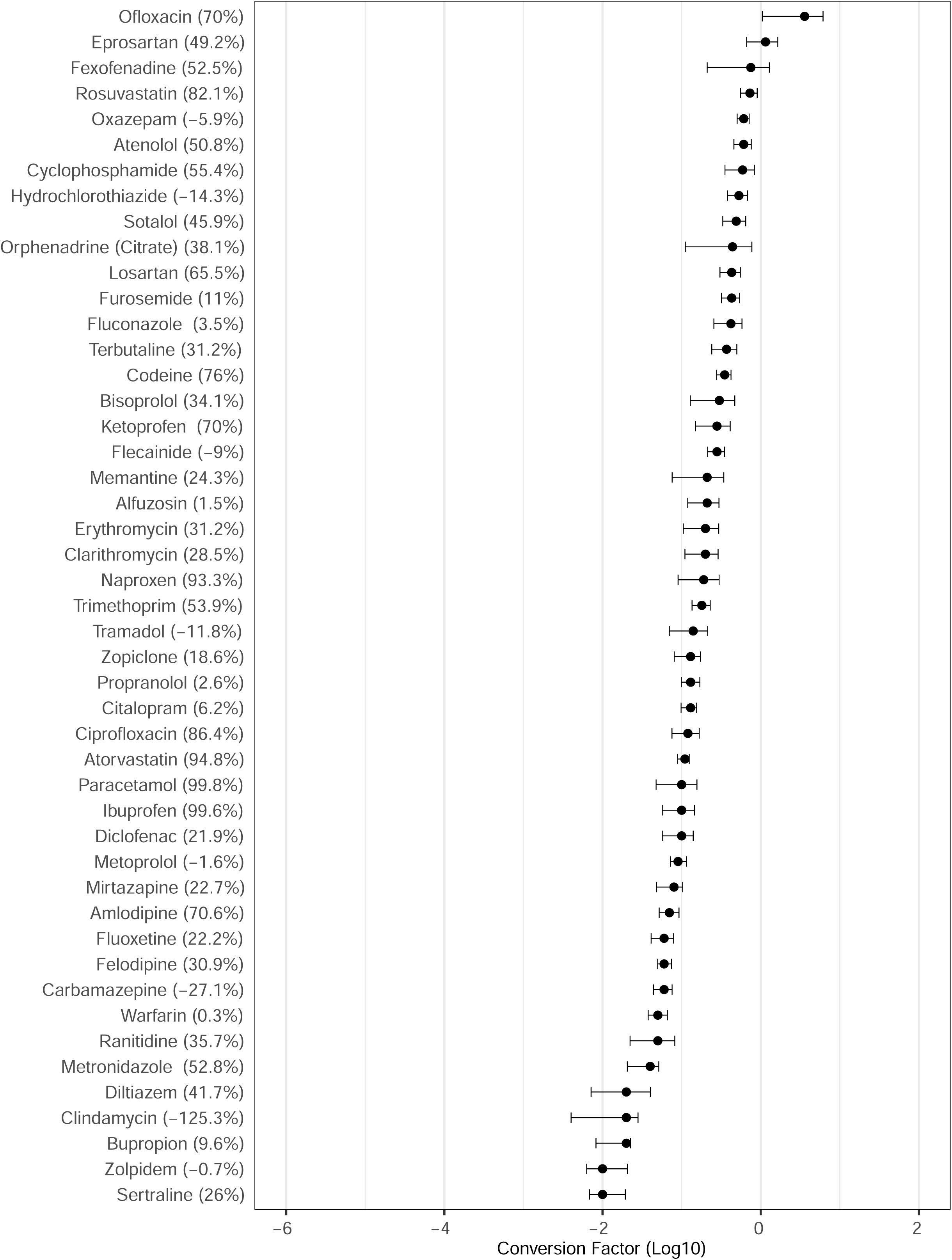
Conversion factor (CF) estimates (proportion recovered in untreated sewage in relation to sales) for 53 active pharmaceutical ingredients (APIs). The data is based on regional sales and recoveries in three wastewater treatment plants in Stockholm 2004-2021. Error bars represent 95% confidence intervals. Estimated removal efficiency for each drug is shown in parenthesis on the y-axis where data is available.

The variability around the conversion factor of each API was estimated using the median absolute deviation (MAD) divided by the median value and expressed as a percentage (MADM, %). Estimates ranged between 34.4% for warfarin to 125.2% for orphenadrine (citrate) (Figure 3). For example, the MADM for warfarin indicates that its usage estimate obtained from back-calculating wastewater concentration measurements will give an average error of approximately 1/3 of the estimated value. MADMs were below 100% (corresponding to a two-fold error) for 43 APIs (81.1%), but not for orphenadrine (citrate), ranitidine, memantine, ibuprofen, ketoprofen, ofloxacin, paracetamol, diltiazem, sertraline, and tetracycline.

**Figure 3.**
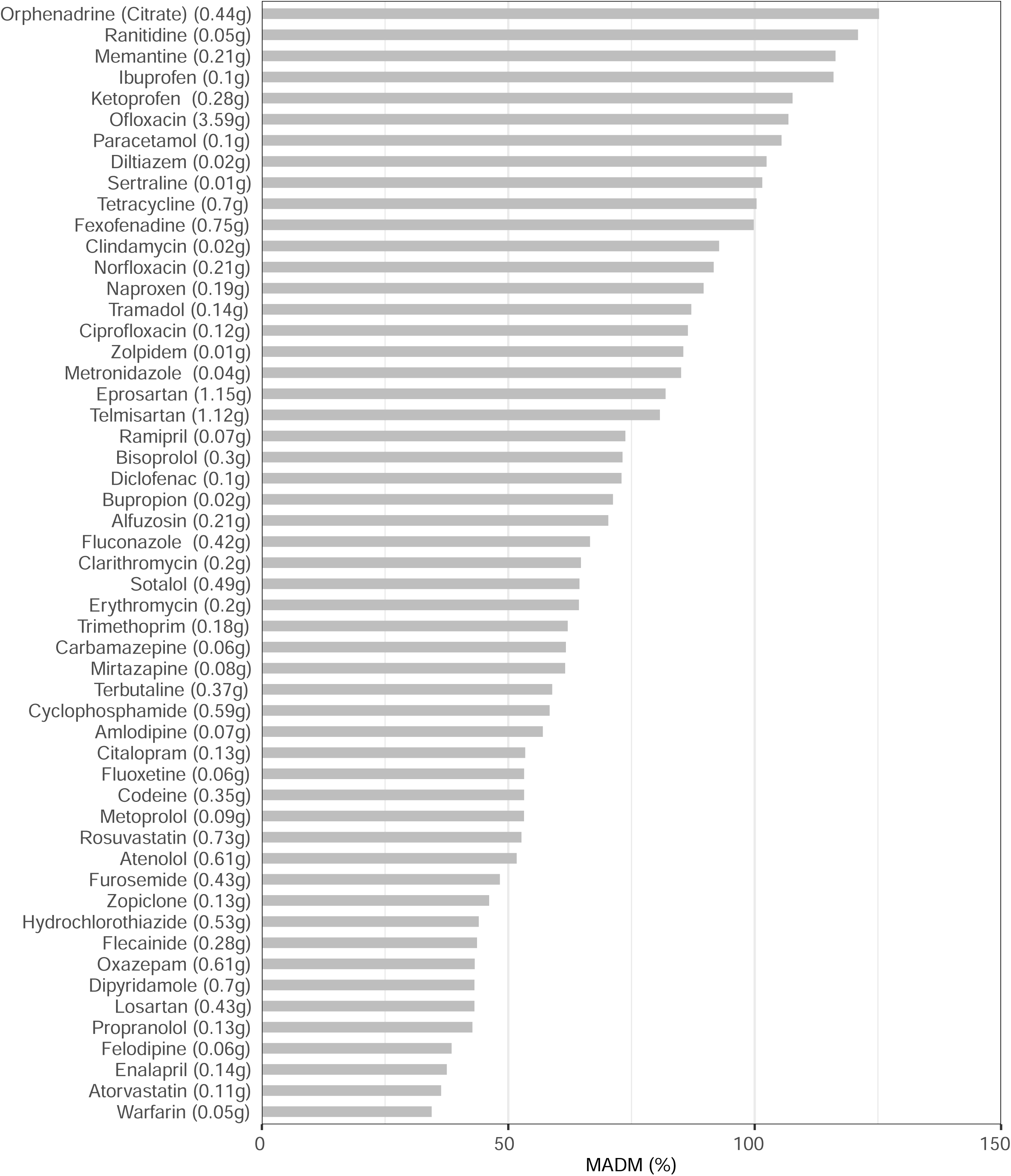
The bars represent the variability of the conversion factors (proportion recovered in untreated sewage in relation to sales). The variability is expressed as Median Absolute Deviation divided by the median values (MADM, %). Median of quantities recovered in wastewater for every gram used are shown in parentheses on the y-axis.

To test the precision of the CFs in estimating API usage, we estimated usage of 53 APIs from mass loads measured in Bromma WWTP between 2004-2021 and compared with the reported usage data. The error in fold change was less than or equal to 2 for 36 APIs (67.9%) (Figure 4). Note that data from Bromma were not used to generate the CFs. There were no systematic differences when we compared CFs generated from each of the three WWTPs.

**Figure 4:**
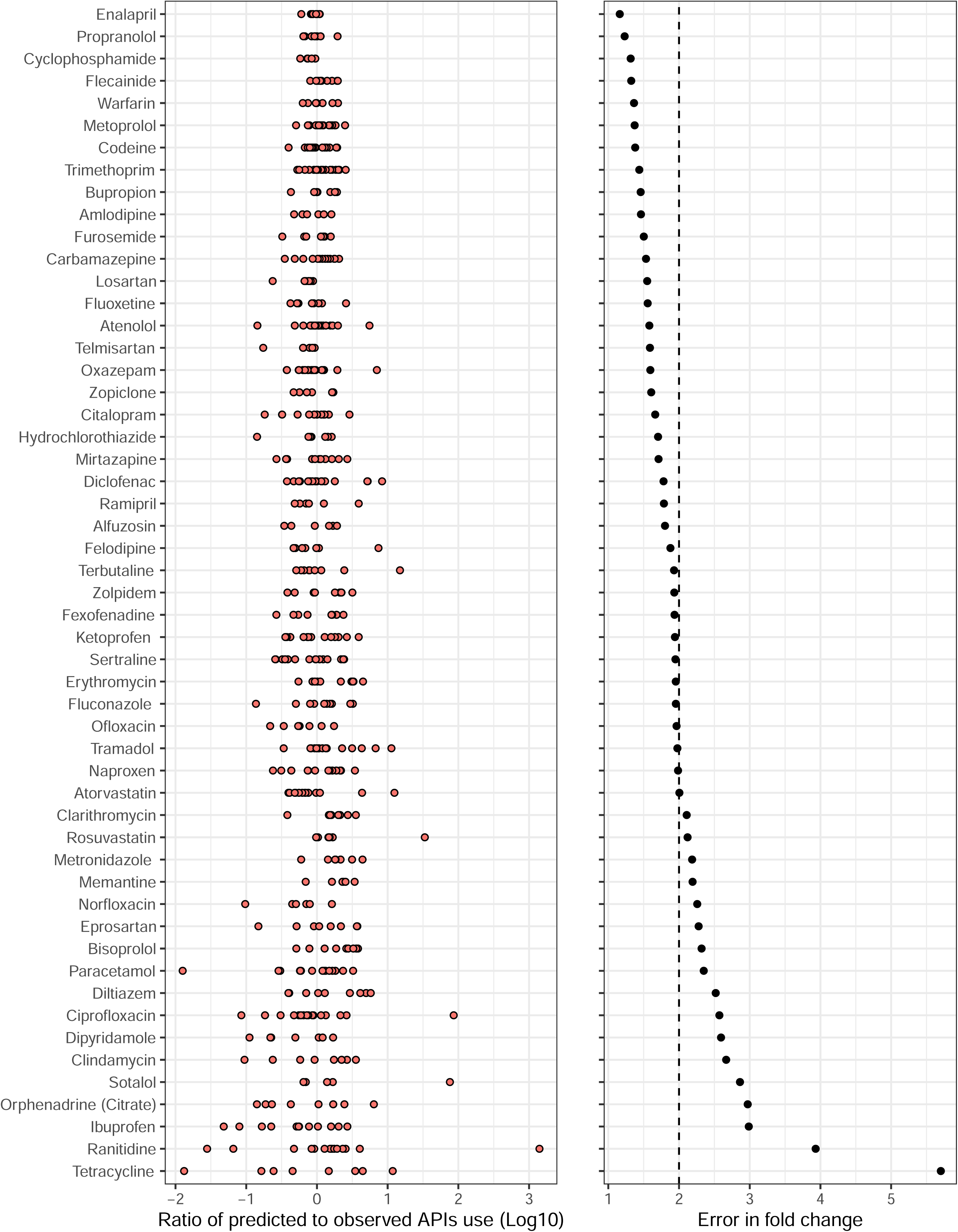
Precision of the conversion factors (proportion recovered in untreated sewage in relation to sales) when predicting usage from active pharmaceutical ingredients’ (APIs) mass loads recovered in Bromma WWTP, Stockholm, Sweden between 2004-2021. The orange dots in the first panel represent the ratios of predicted versus actual reported usage (sales) on a log-scale. The black dots in the second panel represent the error (expressed in fold change units) when predicting usage for each API from wastewater mass loads using the CFs generated from corresponding data at two other WWTPs.

To estimate the removal efficiencies of APIs in wastewater treatment plants (as a potential predictor of CFs and CF-variability), we used the equation ((Influent – Effluent)/Influent) for each year per site where data were available (Supplementary Figure 4). We then used the median estimate as the removal efficiency for each API. Out of the 53 APIs, 47 (88.7%) were included in the removal efficiency analysis for fulfilling the inclusion criterion of having data (influent and effluent concentrations) for at least six years. Our estimates ranged from -125.3% (95%CI; - 315.3 – -50.6) for clindamycin (corresponding to an average increase) to 99.8% reduction (95%CI; 99.4 – 99.9) for paracetamol (Table 1; Figure 2). Low values imply poor API removal by the treatment processes in WWTP, while high values imply excellent removal. For example, 99.8% of paracetamol received in the influents is removed in the effluents. For 8 (17.0%) APIs, including clindamycin, we observed negative removal rates implying higher effluent concentrations than influents (see discussion for possible explanations). Removal efficiency estimates for 39 (83.0%) APIs were positive, implying net removal in the WWTP. We used MADM to estimate the variability of removal efficiency for each API. The variability estimates ranged from MADM of 30.0% for oxazepam to 129.8% for ibuprofen. For 43 (91.5%) APIs, variability estimates were less than 100%.

When we evaluated the correlation between the conversion factors and removal efficiencies using simple linear regression, we observed a weak association (p = 0.04, R^2^=0.07, Supplementary Figure 5). The removal efficiency had, thus, a low influence on the API conversion factors. Neither was lipophilicity a good predictor of CFs (p = 0.11, R^2^=0.03; Supplementary figure 6) nor removal efficiency (p = 0.713, R^2^=0.003; Supplementary figure 7). Since different laboratories measured the API wastewater concentrations between 2004-2011 and 2012-2021, we investigated whether the change in labs affected the CF estimates for 21 of the 53 APIs where data were sufficient for analysis. We observed significant differences between the two periods for 8 of the 21 APIs (Supplementary Figure 8). Out of the 8, drug yield estimates were significantly lower for 7 APIs in 2012-2021 compared with 2004-2011. Next, we compared the variabilities (MADM) of CF estimates between the two periods for these 21 APIs but observed no significant (p=0.103) difference. Lastly, we compared the variability of metoprolol CFs when isotope-labelled standards were used to analyze the wastewater samples or not – data was not available for other APIs. The variability of the CFs estimates from 2019-2021 when the laboratory used isotope-labelled standards for metoprolol detection was 15.5%, which was four times lower than 64.8% (2012-2018) when the corresponding standards were not used.

## 4. Discussion

We have utilized a uniquely long time-series of analyses of APIs at three municipal WWTPs together with corresponding high-quality data on sales to evaluate the potential to estimate the use of medicines from analyses of sewage. We show promising evidence that for the great majority of APIs, usage may be estimated with a precision within a factor of two by back calculations from concentrations measured in the wastewater. Neither lipophilicity nor removal efficiencies appeared to have a large influence on how closely used amounts of APIs are correlated to recovered amounts in wastewater.

For 52 of the 53 APIs, the estimated recovery in wastewater was equal to or less than the estimated use. This is expected given both metabolism in the body as well as potential further metabolism/degradation/sequestration in the sewer systems, thus adding confidence to the CF estimates. However, for ofloxacin the estimated wastewater recovery was significantly higher than the estimated use (3.6 times). Out of all the antibiotics (ciprofloxacin, erythromycin, clarithromycin, clindamycin, metronidazole, norfloxacin, ofloxacin, tetracycline, and trimethoprim) in this study, ofloxacin is the one with lowest use, given through hospital requisitions and prescriptions only, while the remaining antibiotics were additionally sold via open care requisitions and in the case of metronidazole also over the counter. The types of pharmaceutical sales captured in the database we used in this study included all legal routes, that is (i) over the counter sales, (ii) open care requisitions, (iii) prescriptions and (iv) hospital requisitions. We note that the reported wastewater concentrations of ofloxacin were consistently low (average of 28 ng/L, excluding non-detects), potentially contributing to challenges in measurements. Furthermore, it was only reported by the analytical laboratory responsible for the analyses between 2004 and 2012, where we have very limited information on methodology and quality assessment. We therefore suspect that a finding of a CF significantly above 1 for ofloxacin might not be a repeatable finding.

When we used the CFs to predict API usage in a test set, the prediction error (expressed in fold-change units) was less than or equal to 2 for the majority of APIs. This implies that for most APIs, major shifts in usage i.e., above two-fold, can be identified based on wastewater concentration data, but a similar approach may not be very useful when the objective is to detect subtle changes in usage. However, we acknowledge that the test set data (Bromma) in this instance is not completely independent from data of the two sites (Henriksdal and Käppala) used to generate the CFs because; (i) the API concentrations in the wastewater were measured by the same laboratories, and (ii) API usage for each site was estimated by multiplying the total grams sold in the Stockholm region with the respective population proportions. For these reasons, the measurement errors for both wastewater concentrations and API usage could be similar in our training and test sets. Therefore, evaluating the CF estimates on an independent test set would be ideal in testing their robustness.

We compared our findings with those of a recent and completely independent study involving 31 WWTPs in Australia, which reported “correction factors” (the inverse of CFs as used here) for back calculating usage of atenolol, naproxen and carbamazepine from wastewater mass loads (Gao et al., 2021). This was the only study we could find that validated their reported factors with pharmaceutical use data in a different place with similar settings. To compare our findings with theirs, we calculated the reciprocals of their correction factors to obtain CFs of 0.72 (95% CI 0.60-0.85), <0.38 (0.24 – 0.47), and 0.12 (0.10-0.13) for of atenolol, naproxen and carbamazepine, respectively. Their conversion factor for atenolol was highly consistent with our estimated CF 0.609 (95% CI 0.458 - 0.76), while our estimates for naproxen 0.194 (0.09 - 0.298) and carbamazepine 0.06 (0.044 - 0.076) were about two-fold lower. Given the numerous differences between studies, our overall impression is that this represents a relatively good match between generated CFs. The small differences between these estimates might be a reflection of how differences in populations studied can impact the conversion factors. For instance, unlike Sweden, where 1-2% of respondents indicated they would flush unused medicine (Castensson and Ekedahl, 2010), 25% of respondents in a similar Australian study on medicine disposal practices admitted to flushing unused medicines (Bettington et al., 2018). Therefore, it is likely that a higher fraction of dispensed APIs is recovered in wastewater from a population more prone to flushing medicines than not.

Another possible explanation for the observed differences from the Australian study could relate to the chemical analyses. It should be noted that the wastewater chemical analyses in our study were not standardized over the study period as in Gao et al (2021). Therefore, we believe that more precise estimates could be obtained with proper standardization of chemical analysis methods. This notion is further supported by our observation of significant differences between CFs for the same APIs while using wastewater concentration data generated by different laboratories, and by the lower variability of CFs for metoprolol after introducing a dedicated isotope labelled standard. Chemical analyses quality/accuracy is potentially a large source of (systematic) error as observed in the variability of estimates between the two laboratories. Further, the variability observed within the same API and laboratory (and the same year, but between WWTPs) suggests that chemical analysis is also a large source of more (random) errors. We are also oblivious that other sources besides testing laboratories, such as changes in the sewer systems over the years and other environmental changes, could contribute to the observed difference given the difference in periods (2004-2011 versus 2012-2021). With data spanning over 18 years, we opted to use the median absolute deviation divided by the median (MADM) value as a measure of variability around the CF estimates to avoid the influence of outliers. Despite the diverse composition of APIs in our study, we still observed variability of less than two-fold for most of them. Again, given the unstandardized chemical analysis methods in our study and the complexity of the wastewater matrix, this result shows promise in the possibility of estimating usage from wastewater.

The positive correlation between usage and the quantities recovered in wastewater for the majority of APIs was unsurprising. It highlighted the opportunity of using wastewater as a potential source of monitoring usage trends for various pharmaceutical drugs, although variability may still prevent reliable detection of small to moderate changes in sales. Previous studies have reported a similar positive relationship between drug (licit and illicit) consumption estimated in other ways and quantities recovered in wastewater (Baker et al., 2014; Gao et al., 2021).

Removal efficiency as a proxy for stability of APIs in wastewater, was not a good predictor of the CFs. Also, for some APIs we found negative removal efficiencies. The latter may feel intuitively incorrect, but it has been documented earlier for many pharmaceuticals included here e.g. carbamazepine, metoprolol, and tramadol, in different parts of the world (Blair et al., 2015; Gros et al., 2010; Kumar et al., 2022). A common explanation for this phenomenon can be biotransformation within the WWTP, for example the removal of glucuronides from conjugated parent compounds (Jelic et al., 2011; Larsson et al., 1999) or other forms of re-transformation of partly metabolized APIs to the mother substance. Other possibilities include errors in the analysis processes that may have to do with e.g., larger matrix effects in influents compared to effluents (Majewsky et al., 2011). Although we hypothesize that the lipophilicity of the APIs could be important predictors of the CF and stability (removal efficiency) in the wastewater, the failure to establish significant associations points to the presence of other factors besides lipophilicity that could influence these parameters.

While uncertainties remain in all studies including this one, many are likely captured within the variability estimates (MADM) presented here. In our study, the pharmaceutical sales data was available in monthly resolution, and daily usage was obtained by dividing it by the number of days in August and September, which could introduce some level of uncertainty into the estimates of the daily use. However, as the catchment populations are reasonably large, most studied APIs are used by many people every day. Hence, one would expect a rather even use across days within a defined period of the year, limiting the impact of this cause of uncertainty. Some drugs sold in the two studied months will not be used until later, but in a compensatory manner, some drugs purchased before August were most likely used during the sampling period. Similarly, some medicines sold in Stockholm may be consumed when people are elsewhere, and vice versa. For reference, Stockholm registered about 1 million nights spent at tourist accommodation establishments in August 2021 (Eurostat, 2021), corresponding to 1-2% of the residents. The largest consumers of medicines in society are also the very old, who may not be travelling as much as younger people. Taken together, we therefore believe travel had a relatively minor net effect in this case. Notably, travel also would only affect those medicines sold in pharmacies and shops but not those administered within hospitals to inpatients.

Sweden has an existing recommendation established already in 1971 of returning unused drugs to the pharmacies for destruction rather than disposing of them in the garbage or flushing them into the sewers (Persson et al., 2009). Returns have been estimated to be between 2.3% and 4.6% of the sold amount (Ekedahl et al., 2003). How large the true unused proportion is, however, difficult to conclude, as is where they end up. Given the long-standing recommendation in Sweden as well as international comparisons of questionnaire results (Castensson and Ekedahl, 2010), the Swedish population likely flushes down a smaller proportion of unused drugs than people in many other countries. Flushed medicines would bypass human metabolism and be equivalent to 100% excretion. Hence, an increase in flushing would be expected to increase the mass flow to WWTPs more than a corresponding increase in actual use would do. This would, in turn, mean that applying CFs generated under conditions with low flushing (as is likely the case here), might lead to overestimations of use (at least slightly) in regions with a higher flushing rate. The largest effect would be expected for APIs with extensive human metabolism and corresponding low excretion rate as the parent compound.

Proximity of use to the WWTPs could potentially influence variability in wastewater measurements, as it is at least theoretically possible that more of the used substance may be retrieved in treatment plants located close to the population using the medicines than in WWTPs that are distant from the contributing populations. In this study, we used data from three WWTPs in Stockholm, the most densely populated region in Sweden, hence with many contributors relatively close to the WWTP. On the other hand, the longest sewer pipes in the studied areas are approximately 20 km. The temperature in the sewer systems during the sampling period in Stockholm is usually between 19 and 21°C (Bergstrand, 2020). It remains to be studied how such factors, as well as other wastewater properties, may influence biodegradation and biotransformation of APIs in the sewer systems, and in turn CFs. Besides degradation, there is a potential for leakage out from the sewers, a valid concern in many regions with sewer pipes that are sometimes more than a hundred years old.

Polymorphisms in drug metabolism genes and their expression between populations could also be relevant when applying these estimates for certain APIs. Hatta et al., (2015), for instance, showed that Swedes had significantly higher frequency of Cytochrome P450 isoenzyme CYP2C9 defective variant alleles (*2 and *3) than Koreans; hence, its activity is much lower in the Swedes compared to Koreans. CYP2C9 plays an active role in the metabolism of several therapeutically important medicines, including oral anti-coagulants like warfarin (He et al., 2011). Since age- and gender-related differences have been reported in human pharmacokinetic studies (O’Malley et al., 1971; Sotaniemi et al., 1997), we acknowledge that CFs might vary in settings with a different population structure.

Lastly, the complex nature of the sewage matrix poses an analytical challenge in reliably estimating the concentrations of different APIs present (van Nuijs et al., 2015). The most common choice of analytical method for measuring pharmaceutical concentrations in wastewater is liquid chromatography coupled to tandem mass spectrometry (LC-MS/MS), which was also the method used in this study (Fabregat-Safont et al., 2023; Zhang et al., 2021). Still, the use of isotope-labelled standards for every analyte and standardization of all steps would add confidence to measured concentrations and probably contribute to lower variability. In this study, we observed a four-fold reduction in CFs variability for metoprolol when an isotope-labelled standard was used by a testing laboratory to quantify the levels in wastewater samples.

Despite the listed uncertainties and unknowns above, we still observed fairly stable ratios between usage and API recovery in wastewater for the great majority of the studied APIs, even in a non-optimally designed study like ours, where data was not generated for this explicit purpose. We think the results presented here could provide an avenue to begin estimations of medicinal use in resource-strained settings with limited structures, resources or opportunities to track usage of medicines. It may also provide an opportunity to corroborate reported data on sales/usage generated through other means, given that the reliability of such data is sometimes low. We demonstrate that most of the observed usage values fall within a two-fold precision of predicted usage, but that much higher precision was not achieved for the majority of APIs. Therefore, we caution that back calculations from wastewater may not be useful in detecting subtle changes in usage. A remaining limitation of its applicability is that it requires an existing sewage collection system that enables sampling, but notably not sewage treatment infrastructure. Given variability in conditions expected across the world, more studies that establish CF in different regions would add further confidence to the application of the generated CFs in new settings.

## Data Availability

All data produced in the present study are available upon reasonable request to the authors.

## Acknowledgements

We would like to thank Helena Ramström and Annika Hahlin from Region Stockholm for compiling and providing the pharmaceutical sales data used as a proxy for usage in the study. We are also grateful to Johanna Borgendahl from Region Stockholm for providing the wastewater concentrations data of all the APIs investigated and the influent flow-rate data of the wastewater treatment plants on the respective sampling days. Lastly, we thank Gbemisola Allwell-Brown for her contributions in the earlier stages of the study. This work was supported by the SciLifeLab & Wallenberg Data Driven Life Science Program (grant: KAW 2020.0239).

---

**Supplementary Figure 1:**
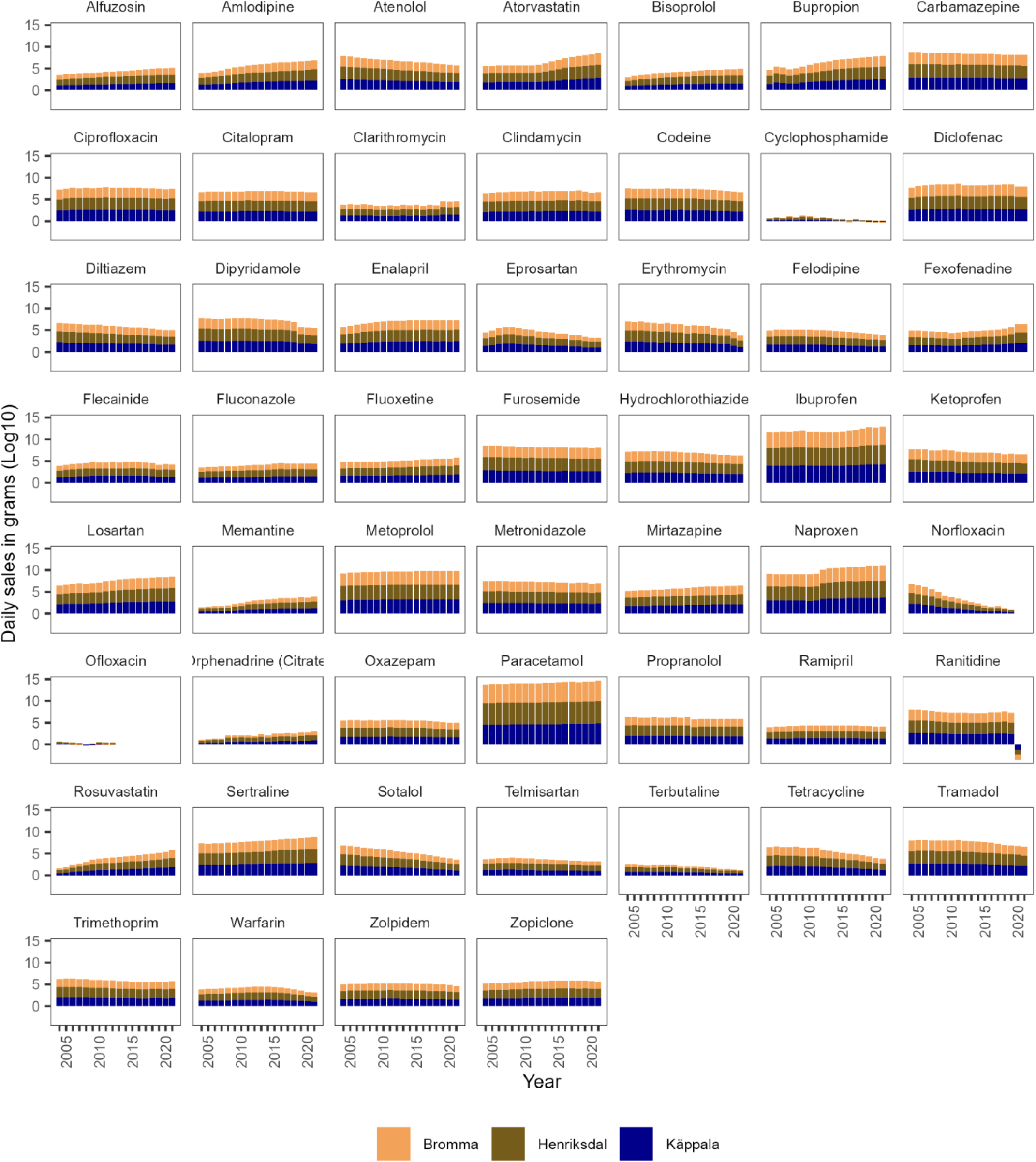
Estimated daily use of 53 active pharmaceutical ingredients (APIs) in catchment areas of 3 wastewater treatment plants (Bromma, Henriksdal, and Käppala) in Stockholm, Sweden between 2004 and 2021.

**Supplementary Figure 2:**
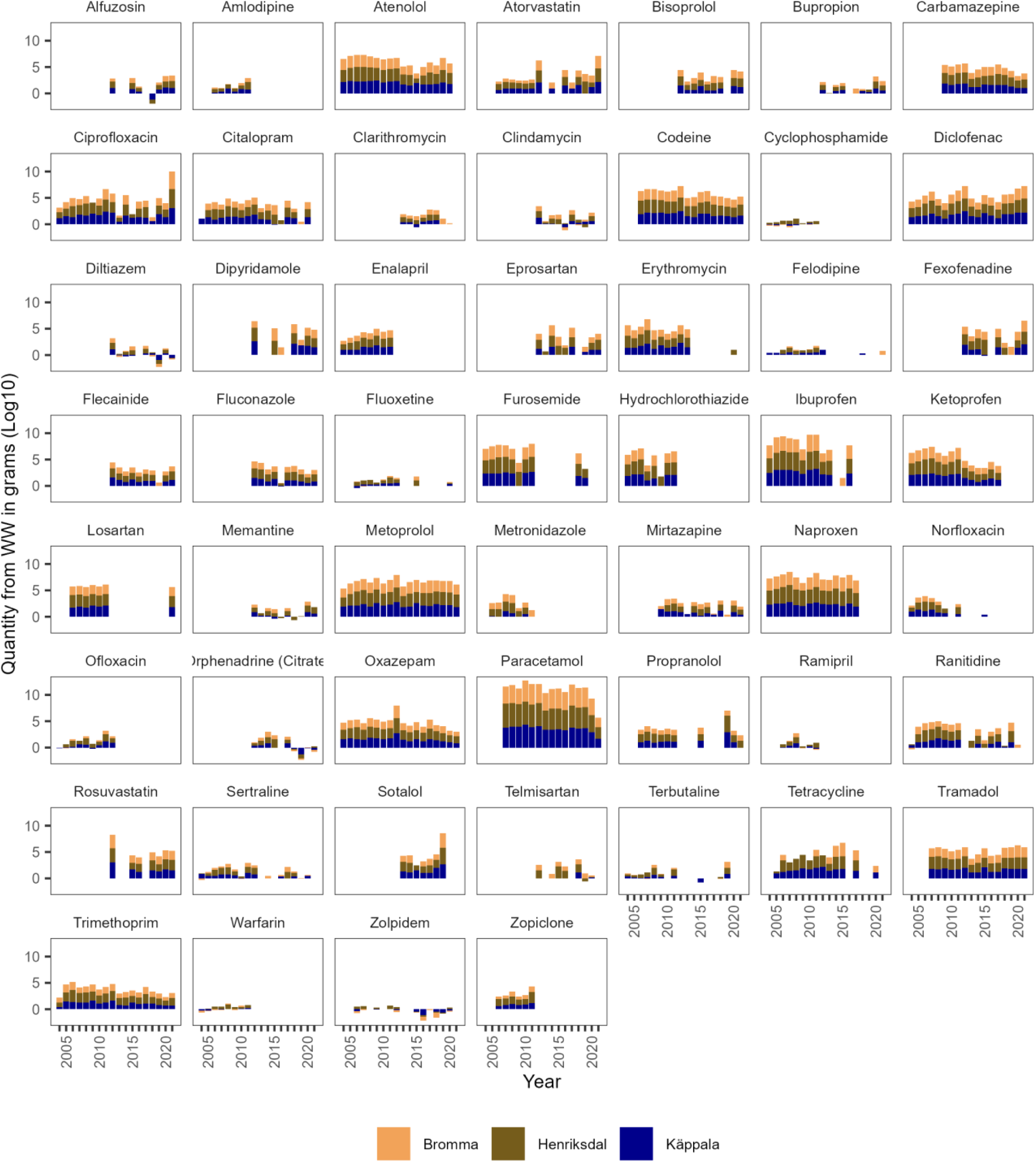
Daily recovered mass loads of 53 active pharmaceutical ingredients (APIs) from 3 wastewater treatment plants in Stockholm, Sweden between 2004 and 2021.

**Supplementary Figure 3 (a-g):**
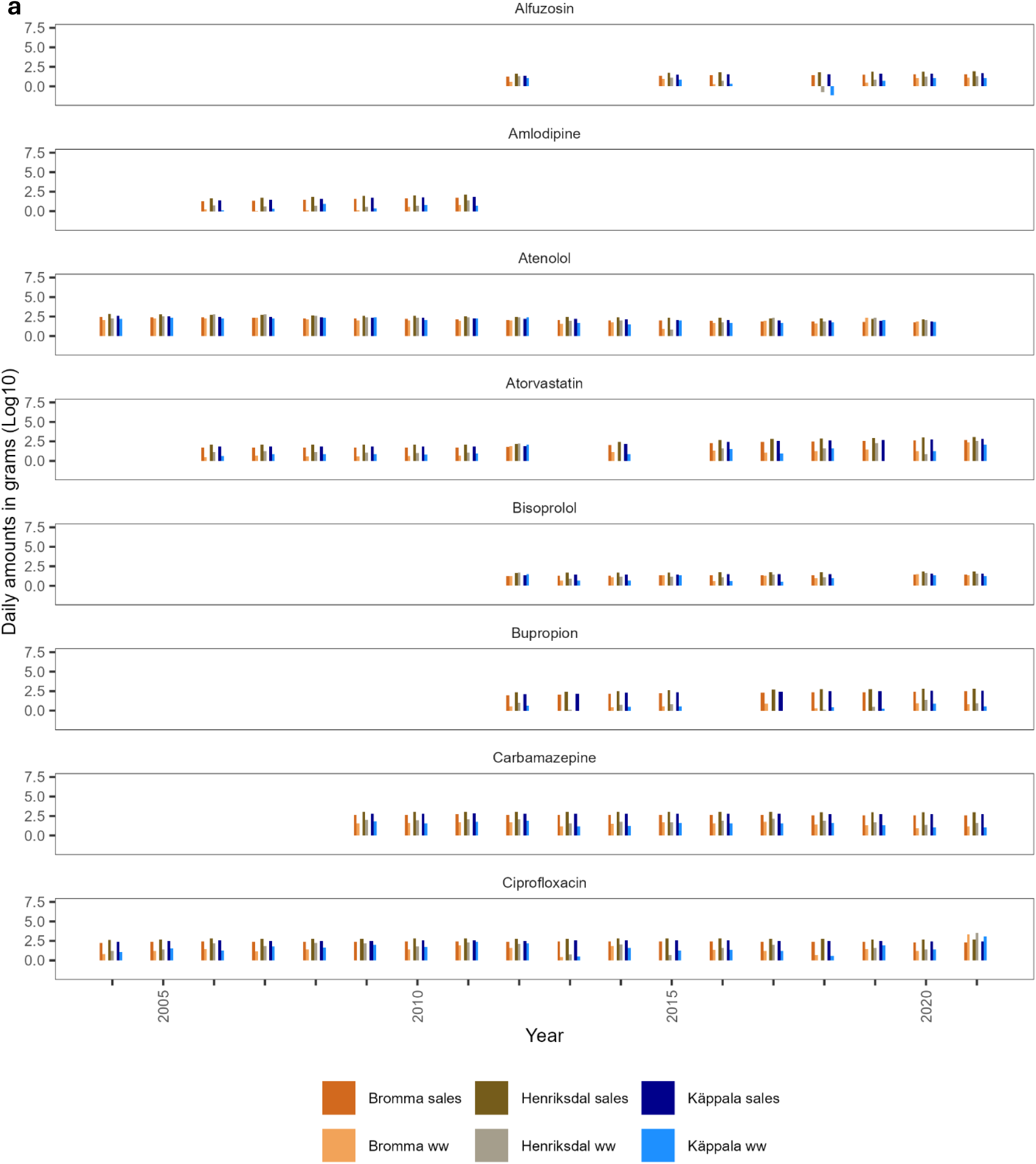
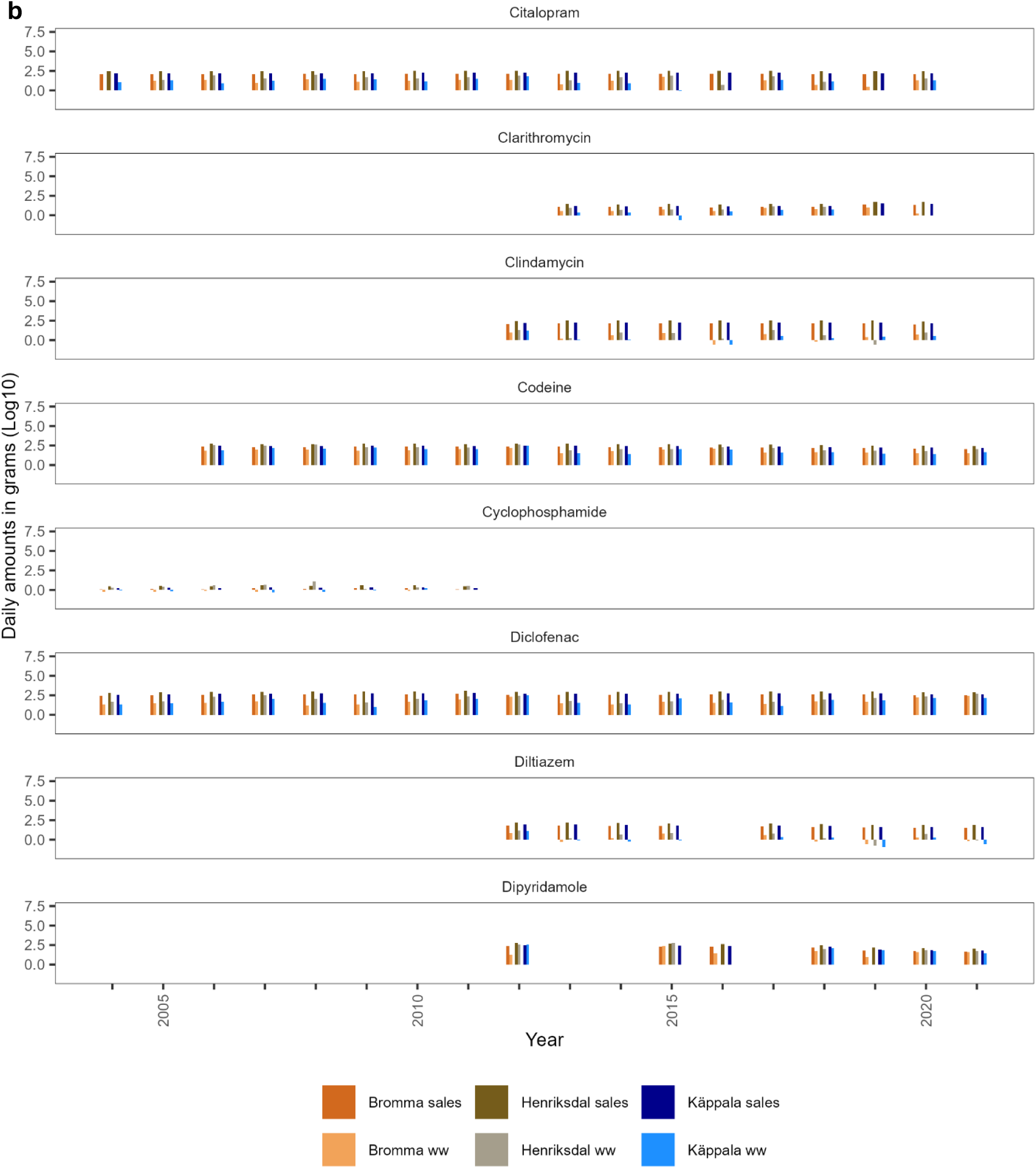
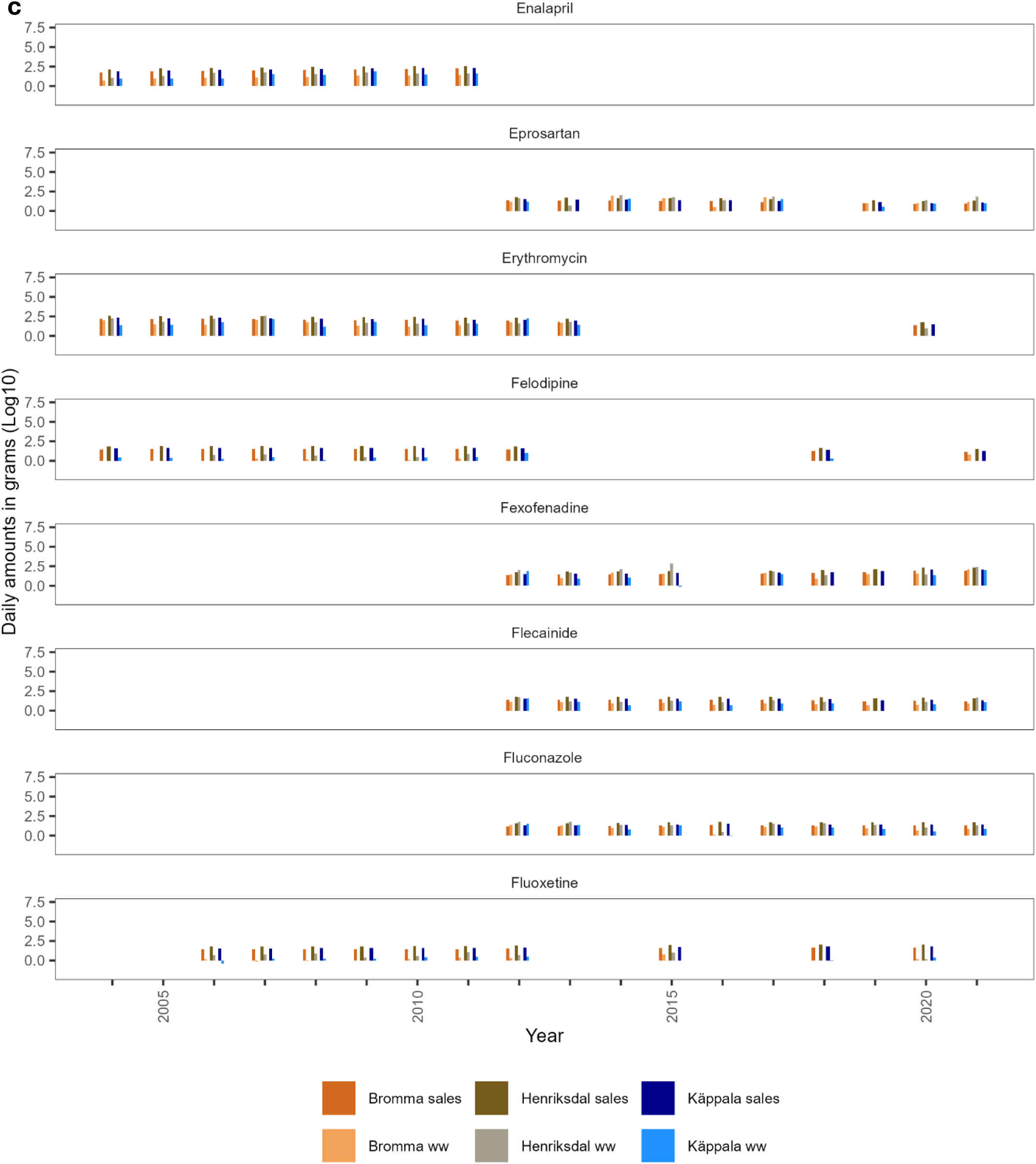
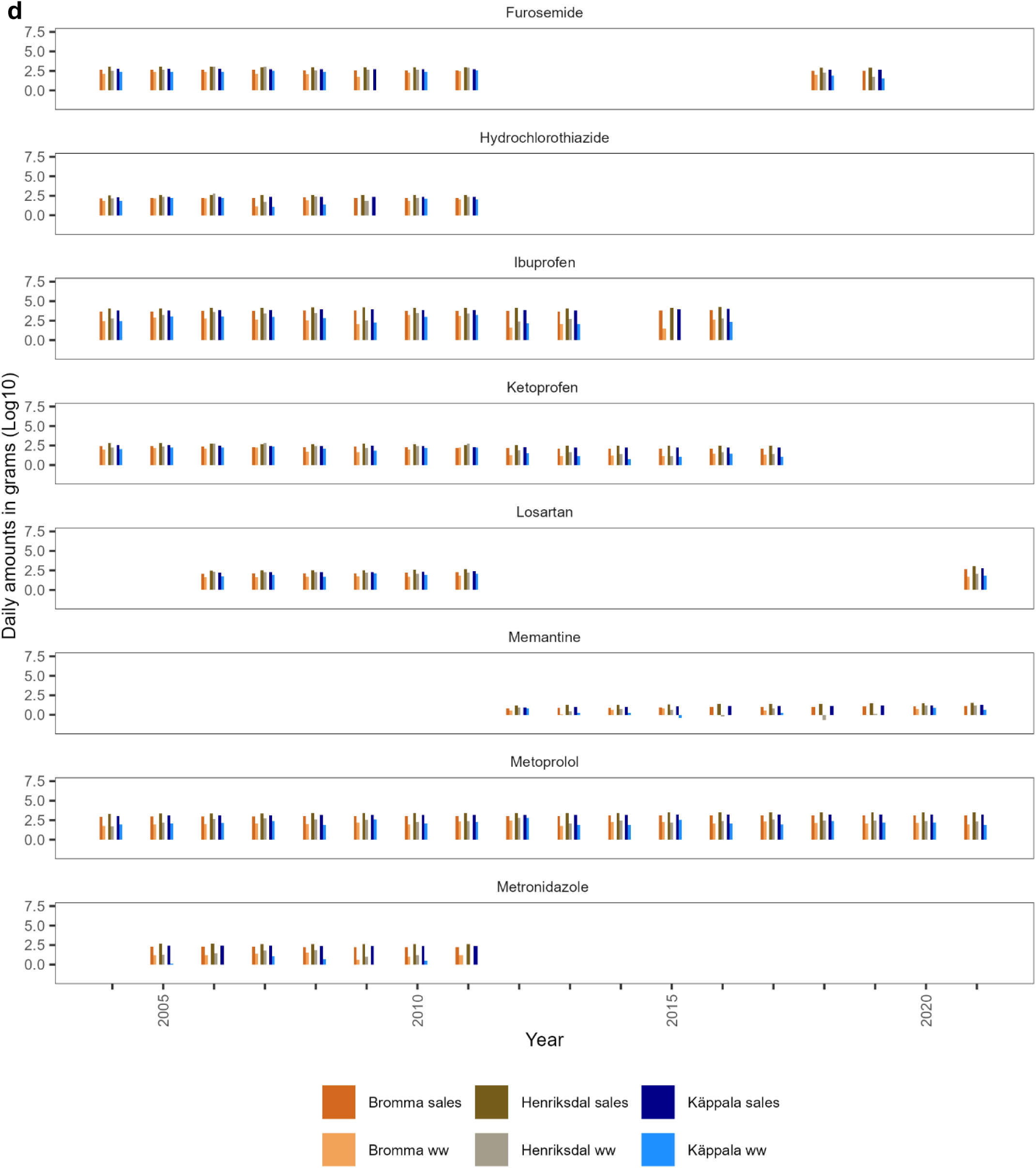
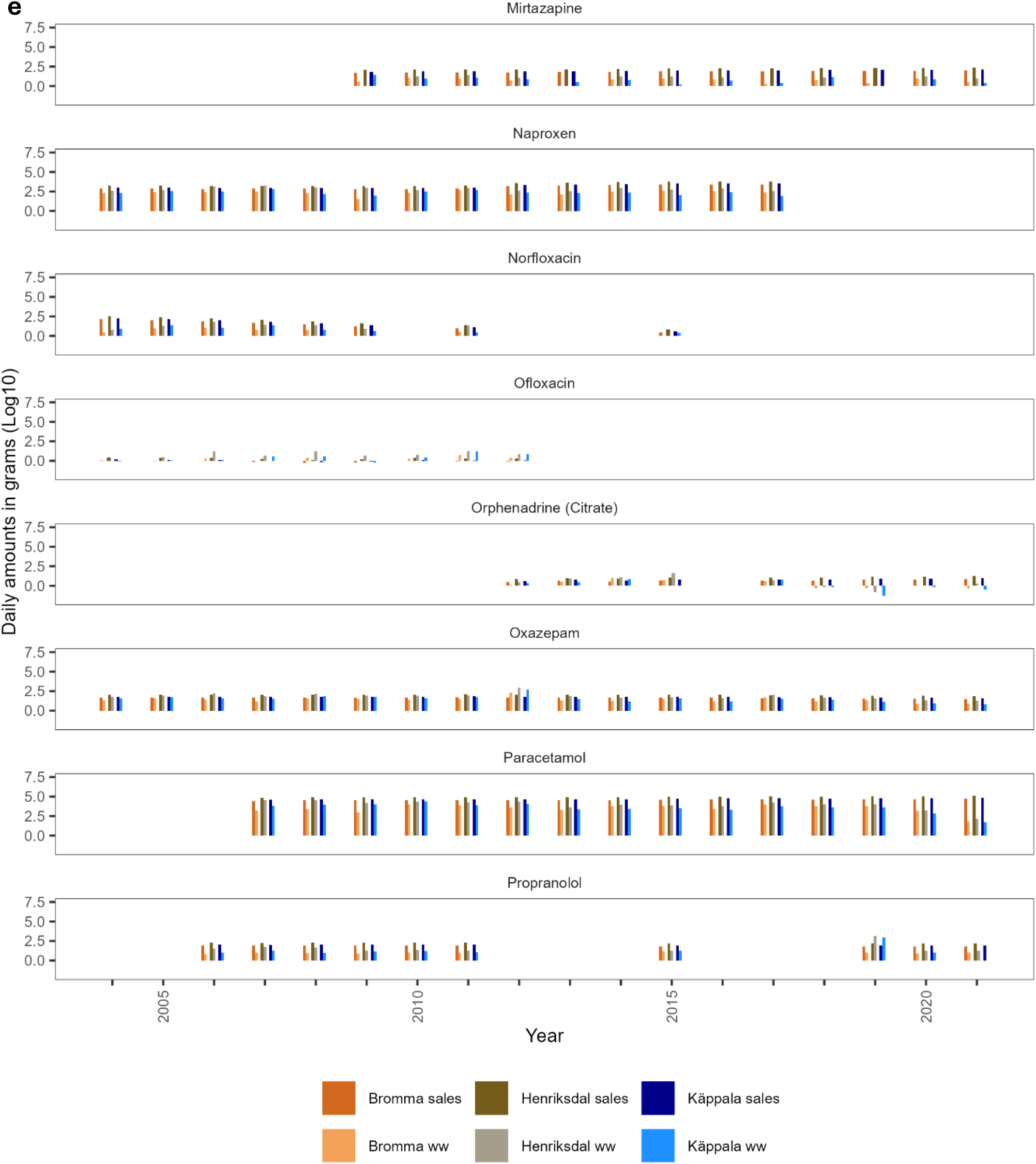
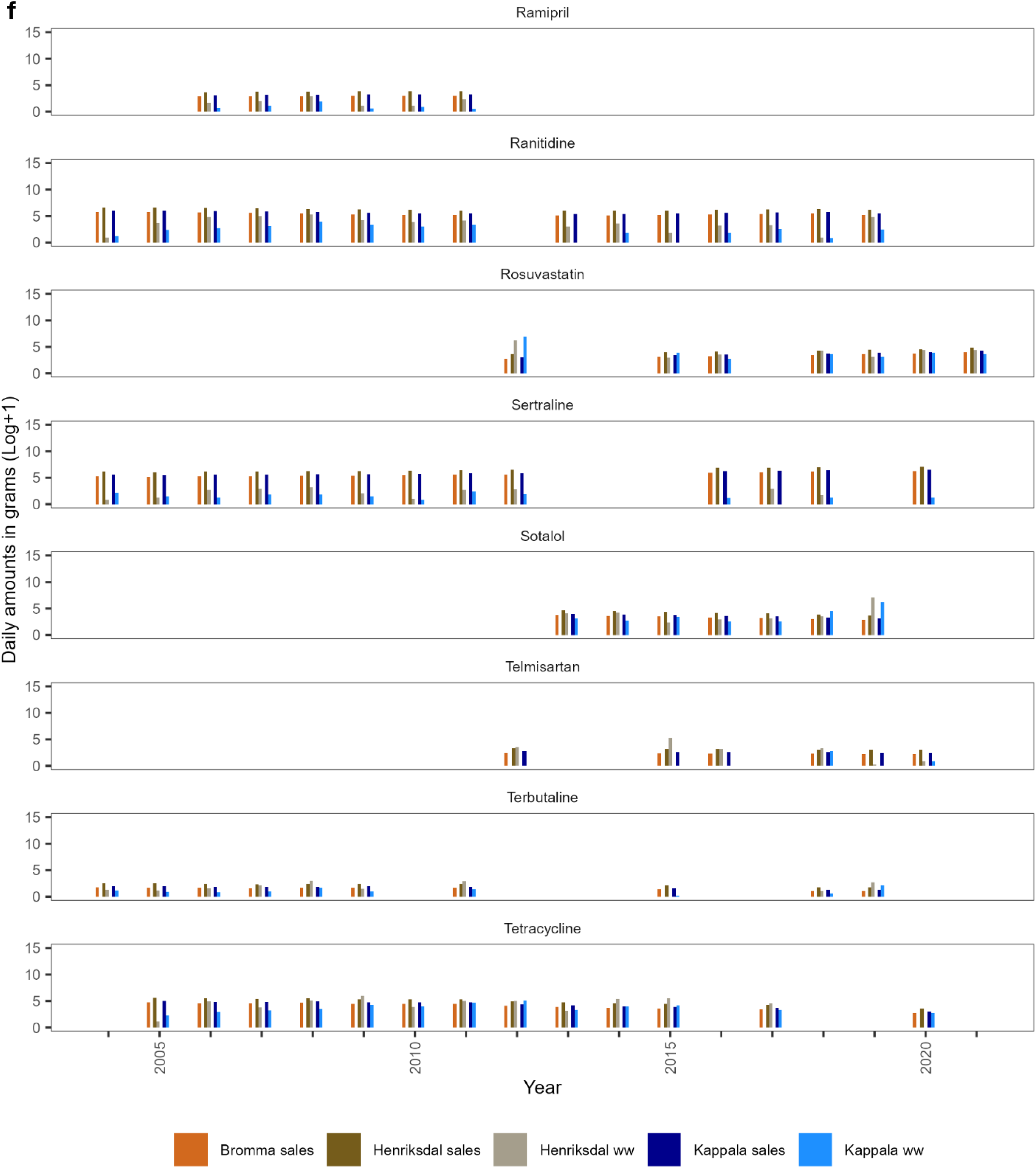
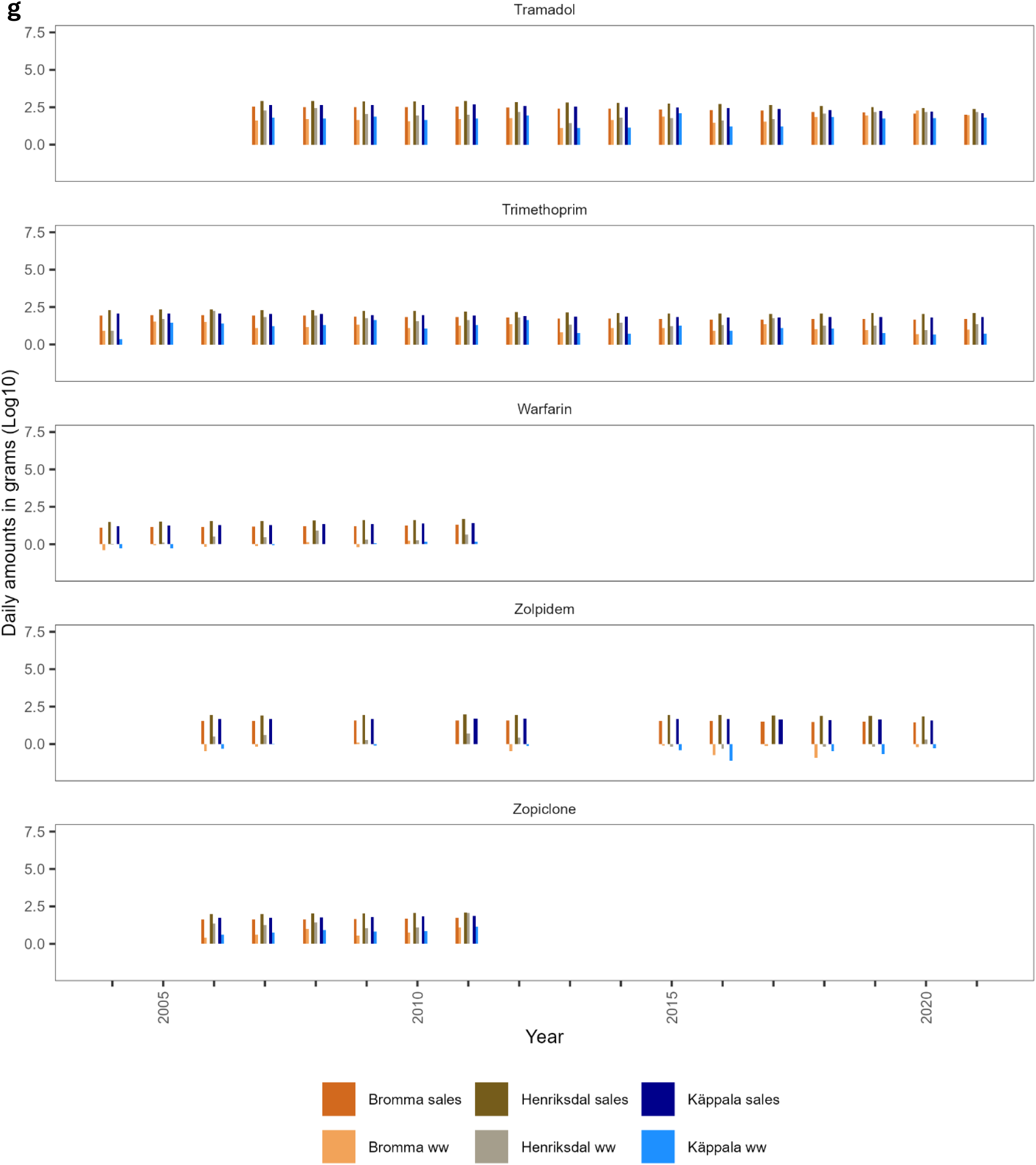
Drug-use changes over the years and the quantities recovered in the untreated wastewater (ww).

**Supplementary Figure 4:**
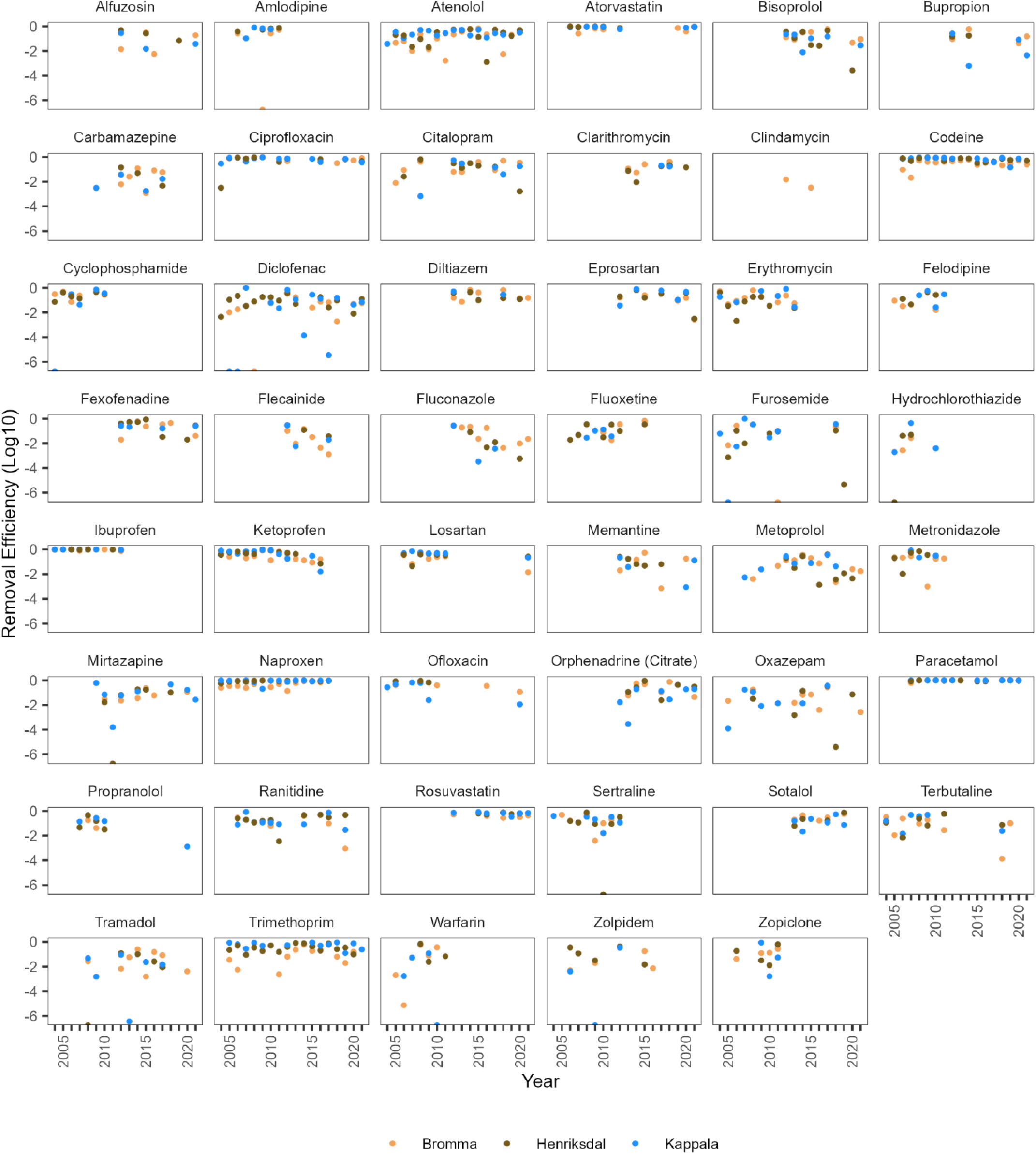
Removal efficiency of active pharmaceutical ingredients (APIs). The dots represent removal efficiency estimates of 47 APIs in 3 wastewater treatment plants between 2004 – 2021.

**Supplementary Figure 5:**
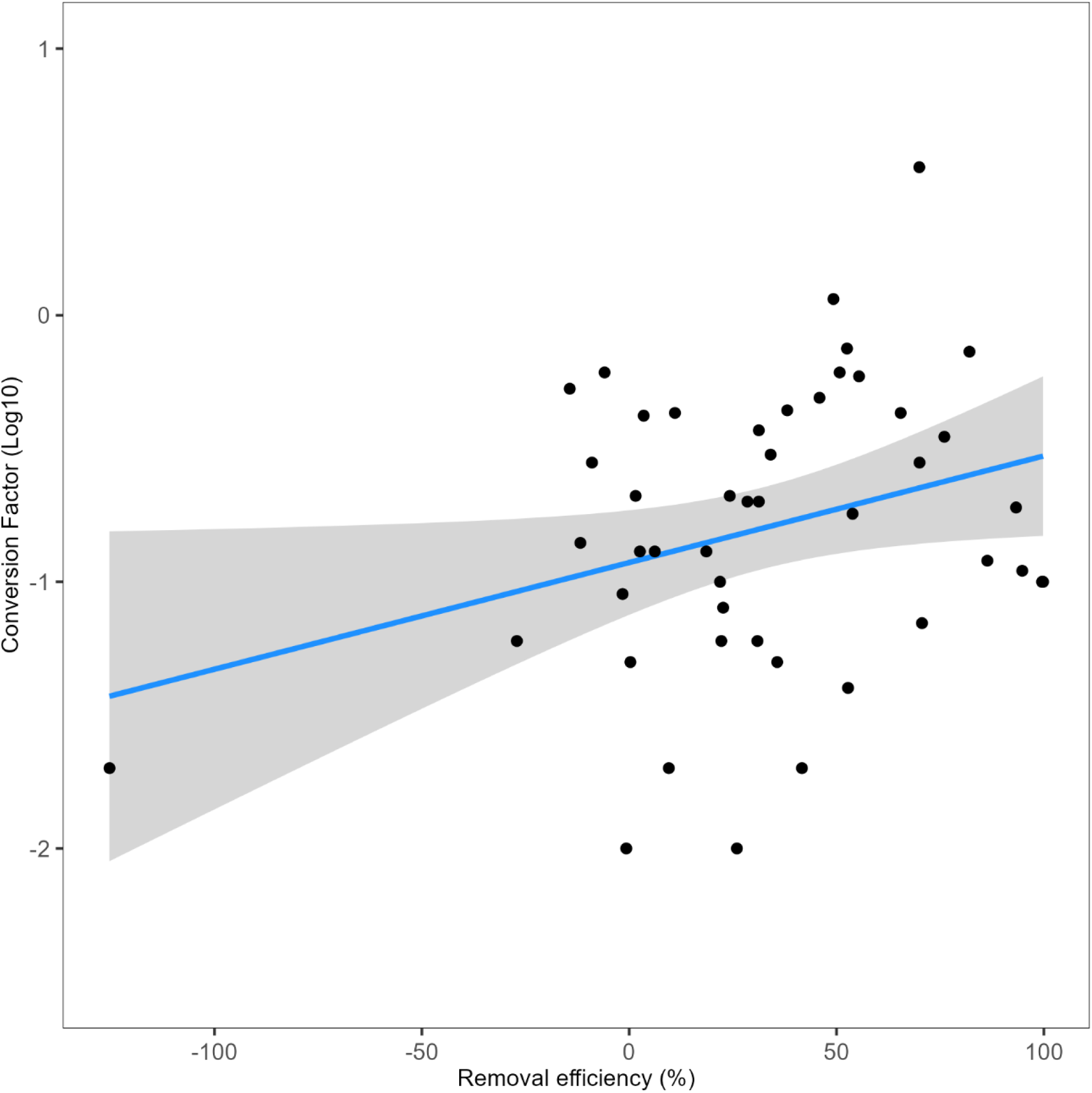
Correlation between conversion factors and removal efficiency. The dots represent individual active pharmaceutical ingredients (APIs), the line represents a significant regression equation (R^2^=0.07, p=0.04), and the shaded area the 95% CI.

**Supplementary Figure 6:**
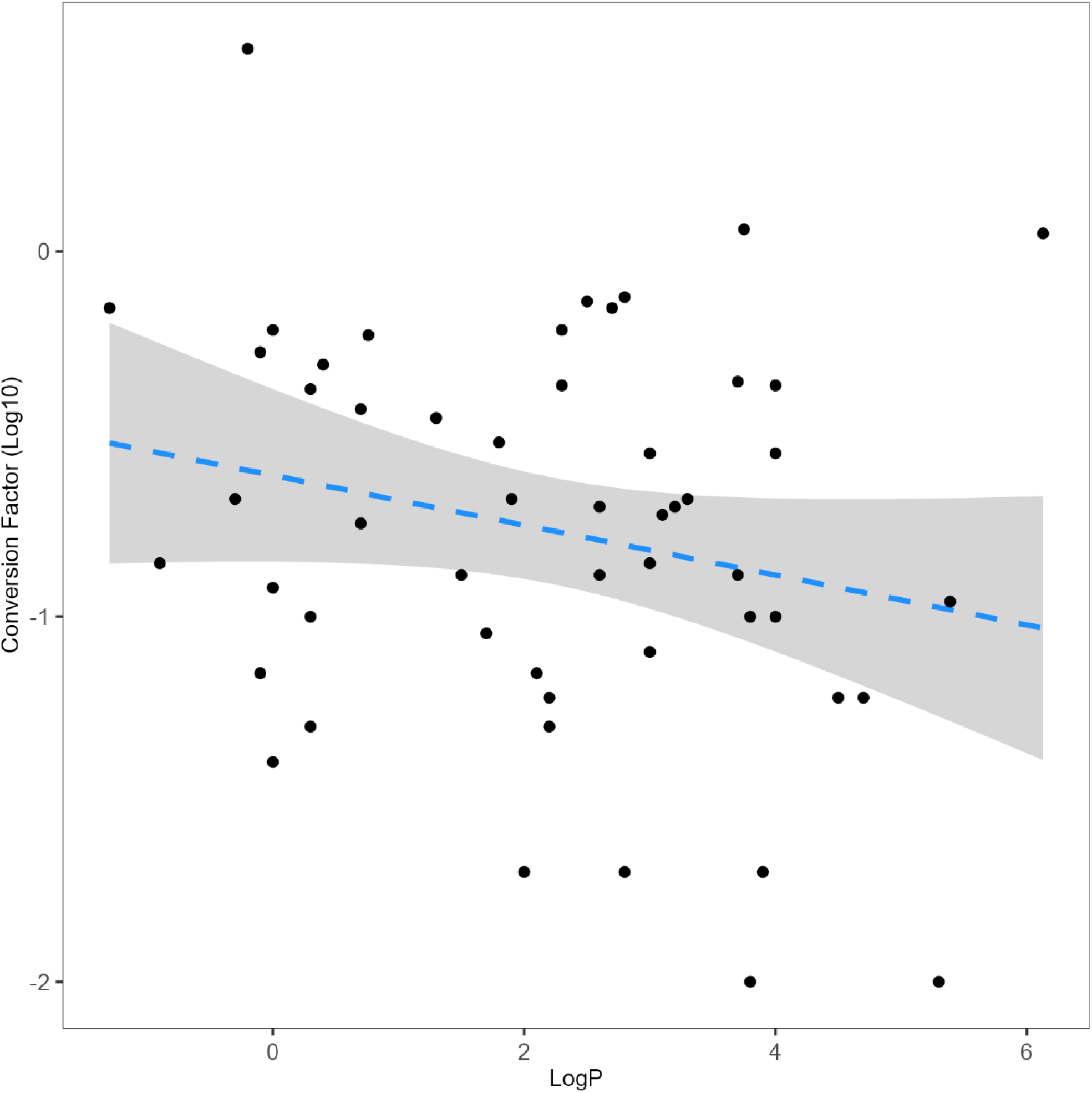
Correlation between conversion factors and lipophilicity (LogP). The dots represent individual active pharmaceutical ingredients (APIs). Dashed line represents a non-significant regression equation (R^2^=0.03, p=0.11), and the shaded area the 95% CI.

**Supplementary Figure 7:**
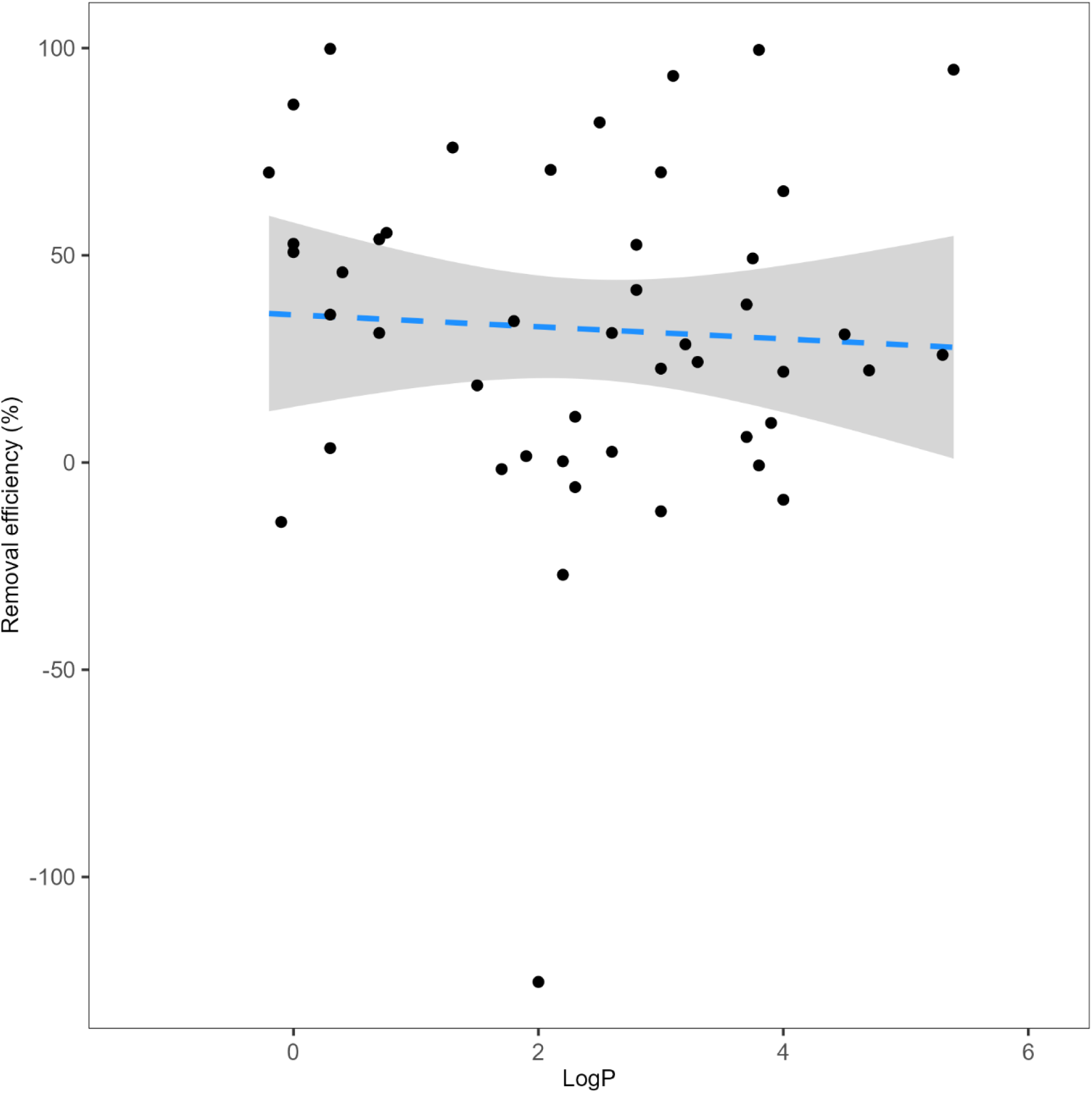
Correlation between removal efficiency and lipophilicity (LogP). The dots represent individual active pharmaceutical ingredients (APIs). Dashed line represents a non-significant regression equation (R^2^=0.003, p=0.71), and the shaded area the 95% CI.

**Supplementary Figure 8:**
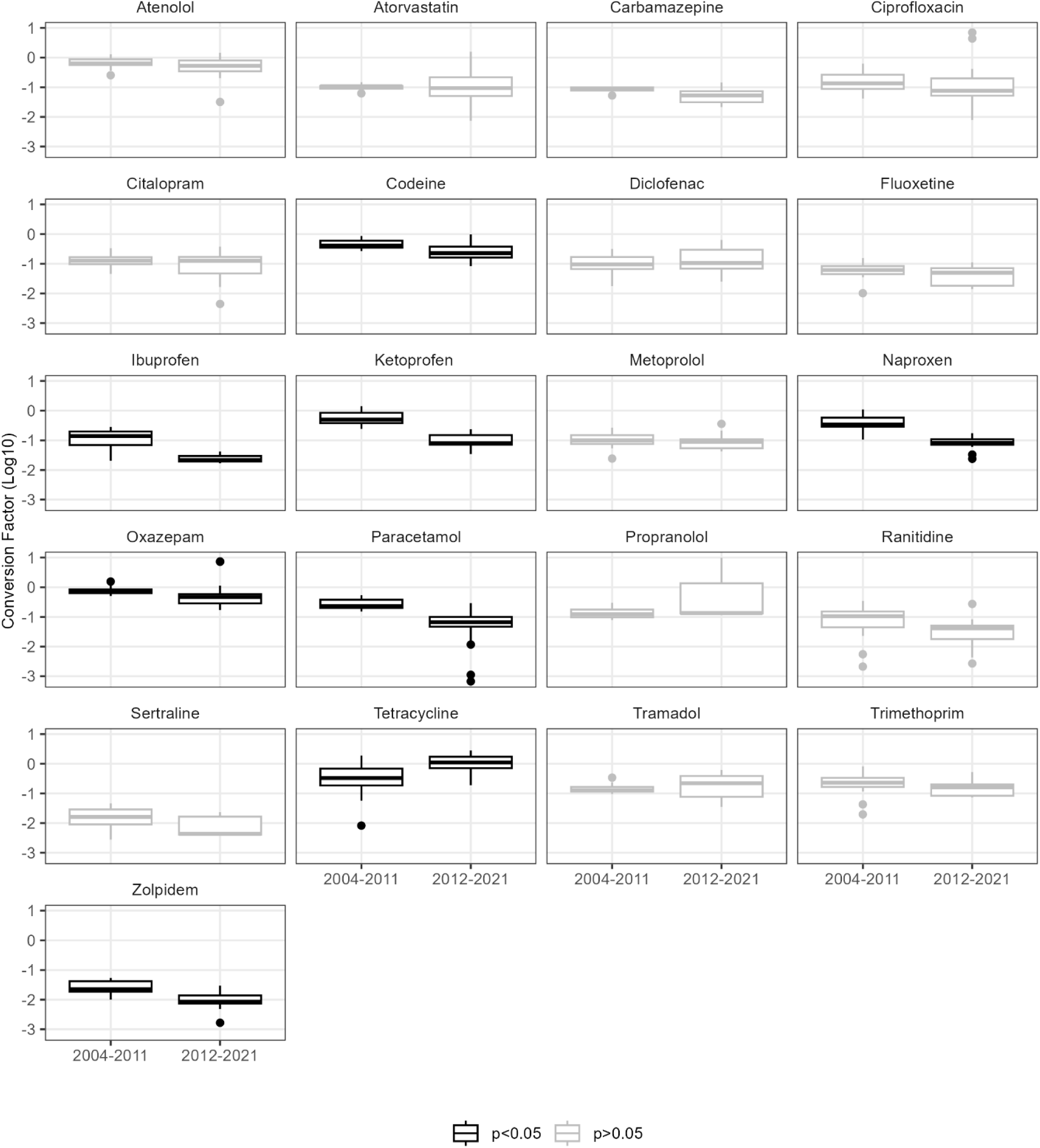
Comparisons of conversion factors (proportion recovered in untreated sewage in relation to sales) between 2004-2011 and 2012-2021, corresponding to a switch in analytical laboratories.

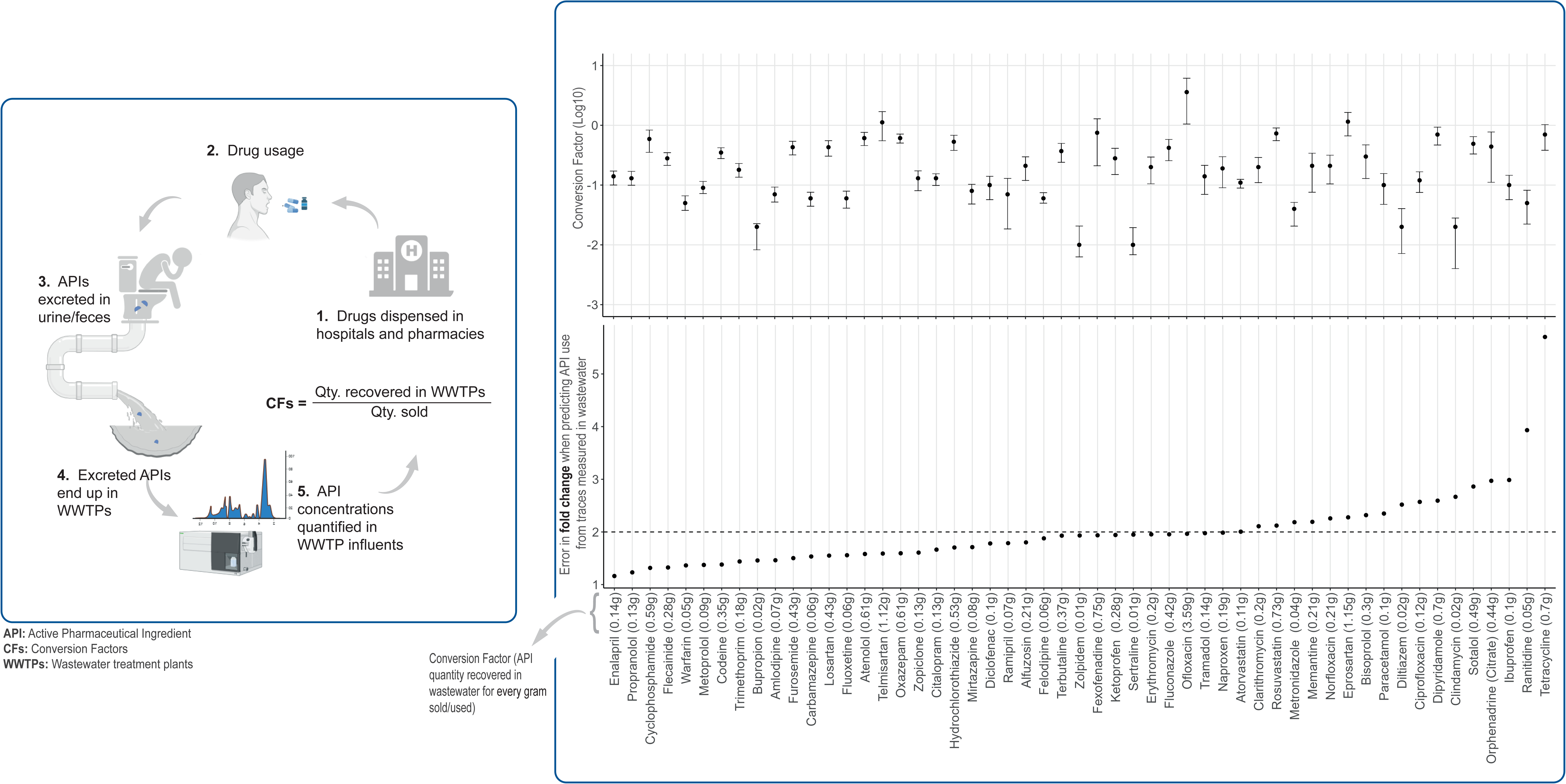

